# Prompts to Table: Specification and Iterative Refinement for Clinical Information Extraction with Large Language Models

**DOI:** 10.1101/2025.02.11.25322107

**Authors:** David Hein, Alana Christie, Michael Holcomb, Bingqing Xie, AJ Jain, Joseph Vento, Neil Rakheja, Ameer Hamza Shakur, Scott Christley, Lindsay G. Cowell, James Brugarolas, Andrew Jamieson, Payal Kapur

## Abstract

Extracting structured data from free-text medical records at scale is laborious, and traditional approaches struggle in complex clinical domains. We present a novel, end-to-end pipeline leveraging large language models (LLMs) for highly accurate information extraction and normalization from unstructured pathology reports, focusing initially on kidney tumors. Our innovation combines flexible prompt templates, the direct production of analysis-ready tabular data, and a rigorous, human-in-the-loop iterative refinement process guided by a comprehensive error ontology. Applying the finalized pipeline to 2,297 kidney tumor reports with pre-existing templated data available for validation yielded a macro-averaged F1 of 0.99 for six kidney tumor subtypes and 0.97 for detecting kidney metastasis. We further demonstrate flexibility with multiple LLM backbones and adaptability to new domains utilizing publicly available breast and prostate cancer reports. Beyond performance metrics or pipeline specifics, we emphasize the critical importance of task definition, interdisciplinary collaboration, and complexity management in LLM-based clinical workflows.

## Introduction

Extracting structured information from free-text electronic medical records (EMR) presents a significant challenge due to their narrative structure, specialized terminology, and inherent variability.^1^ Historically, this process has been labor-intensive and error-prone, requiring manual review by medical professionals,^2–4^ thereby hindering large-scale retrospective studies and real-world evidence generation.^5^ Consequently, automated, reliable methods are needed to extract clinically relevant information from unstructured EMR text.^6^

Natural language processing (NLP) techniques, including rule-based systems and early neural models, have struggled with the nuances of the medical domain.^7,8^ While transformer-based architectures like ClinicalBERT,^9^ GatorTron,^10^ and others,^11–13^ furthered the state-of-the art, they often require extensive fine-tuning on large annotated datasets, which are costly and time-consuming to create.^14,15^ The challenge is particularly acute in specialized tasks like extraction of immunohistochemistry (IHC) results from pathology reports, which requires identifying and mapping tests to the correct results and specimens, resolving synonyms, and navigating diverse terminology.

The rapid emergence of generative large language models (LLMs)^16^ offers a potentially transformative approach. Their large number of parameters and ability to process extensive context windows enable them to retain and ‘reason’ over substantial amounts of domain-specific knowledge without fine-tuning.^17–20^ Natural language prompts allow a high degree of flexibility, enabling rapid iteration and adaptation to new entities and instructions.^21,22^

Recent studies report promising results using LLMs for text-to-text medical information extraction.^23^ Initial efforts have successfully extracted singular/non-compound report-level information, such as patient characteristics from clinical notes,^24^ and tumor descriptors/diagnosis from radiology^25^ and pathology reports.^26,27^ Studies have also demonstrated the potential for extracting inferred conclusions such as classifying radiology findings^28^ and cancer-related symptoms^4^. However, challenges remain, particularly factually incorrect reasoning,^29,30^ and the potential for information loss when forcing complex medical concepts into discrete categories.^31^

Evaluating LLM performance is complicated by the lack of standardized error categorization that accounts for clinical significance and the limitations of traditional metrics like exact match accuracy, which are ill-suited for open-ended generation.^32–36^ For example, misclassifying a test result as “negative” versus “positive” is substantially different than minor grammatical discrepancies between labels e.g. “positive, diffusely” versus “diffuse positive”. This open-ended style thus necessitates methods to constrain generation in order to minimize requisite downstream normalization.^37,38^ Furthermore, many existing clinical NLP datasets utilize BERT-style entity tagging, limiting their use for benchmarking end-to-end information extraction.^33,39,40^ Nonetheless, our lack of pre-annotated data, high degree of entity complexity, and desire for flexibility, coupled with the rapidly improving performance of LLMs,^41^ prompted us to explore their potential.

We present a novel approach to end-to-end information extraction from real-world clinical data that addresses these challenges through three key innovations: flexible prompt templates with a centralized schema for standardized terminology, multi-step LLM interactions with chain-of-thought reasoning, and a comprehensive error ontology developed through iterative ‘human-in-the-loop’^42^ refinement. We demonstrate this approach using renal tumor pathology reports, extracting and normalizing report-level diagnosis, per-subpart/specimen histology, procedure, anatomical site, and detailed multipart IHC results for 30+ assays at both the specimen and tissue-block level. We focus primarily on renal cell carcinoma (RCC) given the high volume of RCC patients treated at UT Southwestern, the diversity of RCC subtypes and wide variety of ancillary studies used for subtyping, and a multidisciplinary UTSW Kidney Cancer Program recognized with a Specialized Program of Research Excellence award from the National Cancer Institute.

First, we detail our pipeline development through the creation of a validated ‘gold-standard’ dataset using 152 kidney tumor pathology reports. Within this we explore our error ontology, used to classify discrepancies based on clinical significance, source (LLM, manual annotation, or insufficient instructions), and crucially, the contexts from which errors arose. We then apply and validate our pipeline using a set of 3,520 institutional kidney tumor reports. Finally, we assess portability using 53 invasive breast cancer and 253 prostate adenocarcinoma pathology reports from an independent, public repository.

Beyond the technical specifics of our pipeline, we place particular emphasis on the broader considerations that arose during development. Specifically, our focus shifted from engineering prompts that *could* extract information, to precisely defining *what* information to extract and *why*. This experience underscores that, as AI approaches human-level intelligence in many domains,^41^ success will increasingly hinge on the clear articulation of objectives, rather than on singular workflow methodologies. As such, by detailing both our success and pitfalls, we hope to provide a roadmap of generalizable context and considerations for future AI-powered clinical information extraction workflows.

## Methods

### Defining the Task

We defined ‘end-to-end’ information extraction as encompassing: (1) entity identification; (2) clinical question inference e.g. what is the *final* diagnosis; (3) terminology normalization; (4) relationship mapping e.g. the IHC result “Positive” corresponds to test “BAP-1” on “specimen A“; and (5) structured output generation. For example, directly converting narrative pathology reports into tabular datasets with columns for specimen/block names, normalized IHC test names, and normalized test results, with each row representing an individual test performed.

Initial entities to extract and normalize from reports included: (1) report-level diagnosis; (2) per-subpart/specimen histology, procedure type, and anatomical site; and (3) detailed specimen and tissue block-level IHC/FISH (fluorescence in situ hybridization) test names and results. We first defined an ‘extraction schema’, outlining standardized labels, a structured vocabulary of terms and preferred synonyms for IHC results, and unique entity specific instructions; see Figure 1A. Unique instructions provided entity-specific context and logic, like differentiating regional vs. distant metastasis based on lymph node involvement for diagnosis.

**Figure 1:**
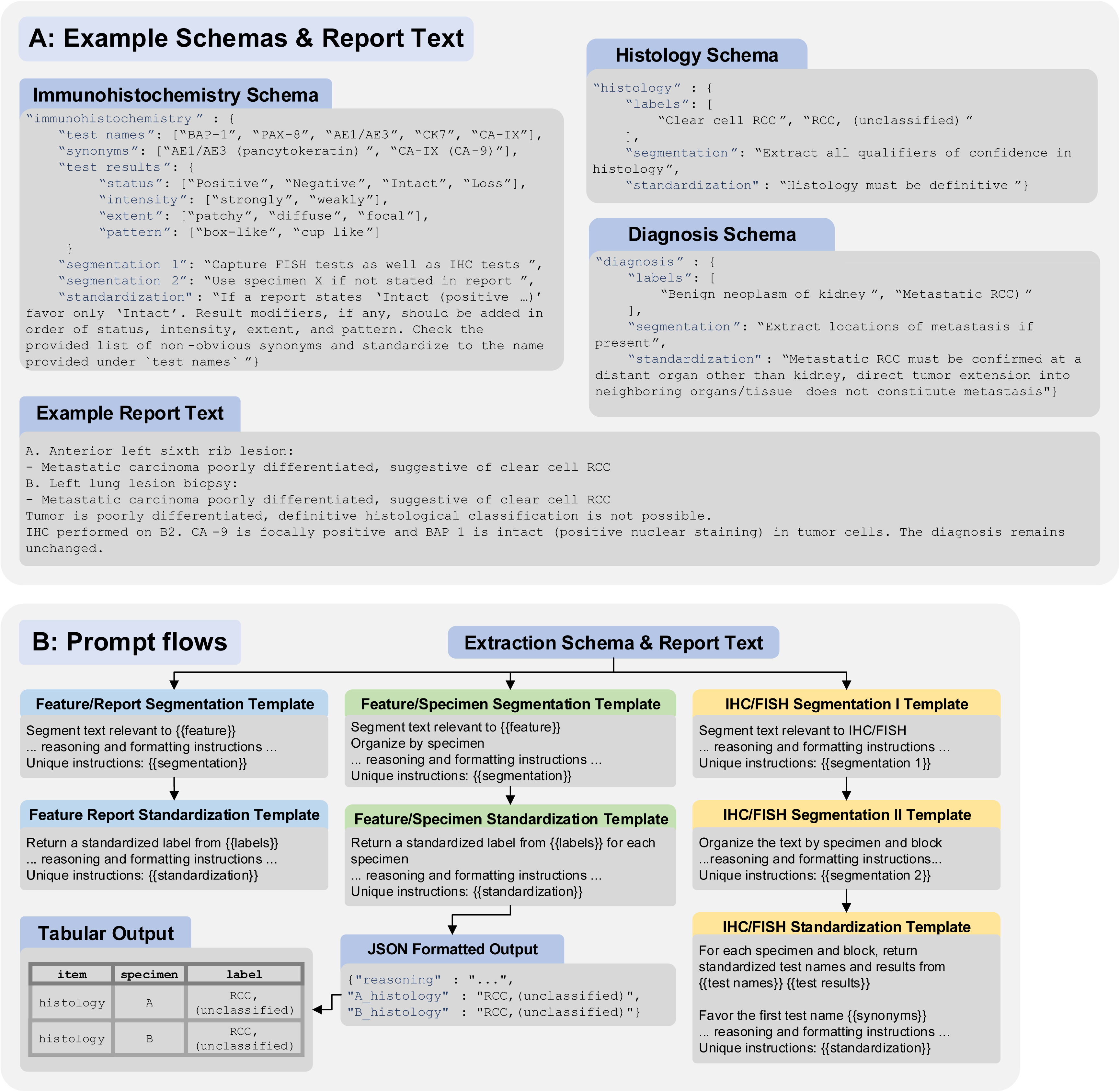
**(A)** Abbreviated examples of the extraction schema for immunohistochemistry (IHC) and fluorescence in-situ hybridization (FISH), histology, and diagnosis, demonstrating the inclusion of entity-specific instructions, standardized labels, and a structured vocabulary for IHC test reporting. An abbreviated report text is included for reference. **(B)** Overview of pipeline steps. Each set included base instructions consistent across entities, and placeholders for entity-specific instructions and labels that could be easily “hot-swapped”, with the {{}} indicating where information from the schema is pasted in. All template sets include initial prompts to segment and organize text, a subsequent standardization prompt to normalize labels and produce structured output, and a final python step for parsing into tabular data. The full output from segmentation steps, both reasoning and the segmented text, is passed to subsequent steps.

Labels for diagnosis were primarily derived from ICD-10 descriptors, with procedure and histology labels primarily sourced from the contemporary College of American Pathologists (CAP) Cancer Protocol Templates for kidney resection and biopsy.^43,44^ Labels for anatomical site and IHC/FISH, along with specialized labels such as the diagnosis “Metastatic RCC” were developed with guidance from pathology experts. While the normalized lists covered common entities, free-text options (e.g., “Other-<fill in as per report>”) were essential for capturing less frequent findings or specific nuances not fully represented, thus preventing information loss.

### Prompt Templates

We used Microsoft Prompt flow^45^ to organize the workflow as a directed acyclic graph, with nodes for Python code execution or LLM requests via specific prompt templates. Reusable, modular prompt templates were designed for portability. We developed three distinct template sets, each optimized for a specific class of entity: The ‘feature report’ set for entities with a single label per report, such as diagnosis; The ‘feature specimen’ set for entities with one label per specimen/subpart; And an IHC/FISH specific set as it uniquely requires matching any number of specimens, blocks, test names, and test results; see Figure 1B. Initial prompts segmented and organized relevant text, while final prompts generated schema-normalized, parsable JSON output.

All prompts required initial ‘reasoning’ generation, passed to subsequent prompts to create a ‘chain-of-thought’.^46^ This has been shown to enhance LLM performance across a wide range of tasks,^47,48^ and provides insight into specific limitations and usage of our instructions.^49^ The reasoning instructions prompted the LLM to consider uncertainties and articulate its decision-making process for text segmentation or standardization. Prompt templates were written in markdown format for organization and included examples of properly structured JSON responses. Full schema and templates are included in the supplement and a GitHub repository for implementation can be found at github.com/DavidHein96/prompts_to_table.

### Workflow Refinement & Gold-Standard Set

We selected 152 reports reflecting diverse clinical contexts from a set of patients with kidney tumor related ICD-10 codes in their EMR history; SFigure1. This included reports with multiple specimens, complex cases that required expert consultation, biopsies and surgical resections, various anatomical sites such as primary kidney and suspected metastases, and both RCC and non-RCC histologies. In total, 89 reports contained local/regional RCC, 41 contained metastatic RCC, 9 contained non-RCC malignancies (such as urothelial carcinoma), and 13 contained benign or neoplasms of uncertain behavior of kidney (such as renal oncocytoma). All data was collected under the purview of our institutional IRB STU 022015-015.

These reports were used to iteratively develop a ‘gold-standard’ annotation set mirroring the target output format. This served to: (1) benchmark extraction accuracy, (2) facilitate iterative ‘human-in-the-loop’^42^ refinement by addressing model-annotation discrepancies, and (3) clarify extraction goals by resolving schema/instruction ambiguities. First, the workflow was executed with preliminary prompts and schema to generate rough tabular outputs, allowing for expedited manual review and reducing annotation burden for creating the initial gold-standard set. Subsequently, the prompts and schema were updated to reflect desired changes, and the workflow was executed again. The output was then matched to gold-standard annotations and discordant results manually reviewed, with source and severity of the discrepancy documented and used to inform subsequent adjustments to the workflow and gold-standard. This process was then repeated iteratively, as outlined in Figure 2.

**Figure 2:**
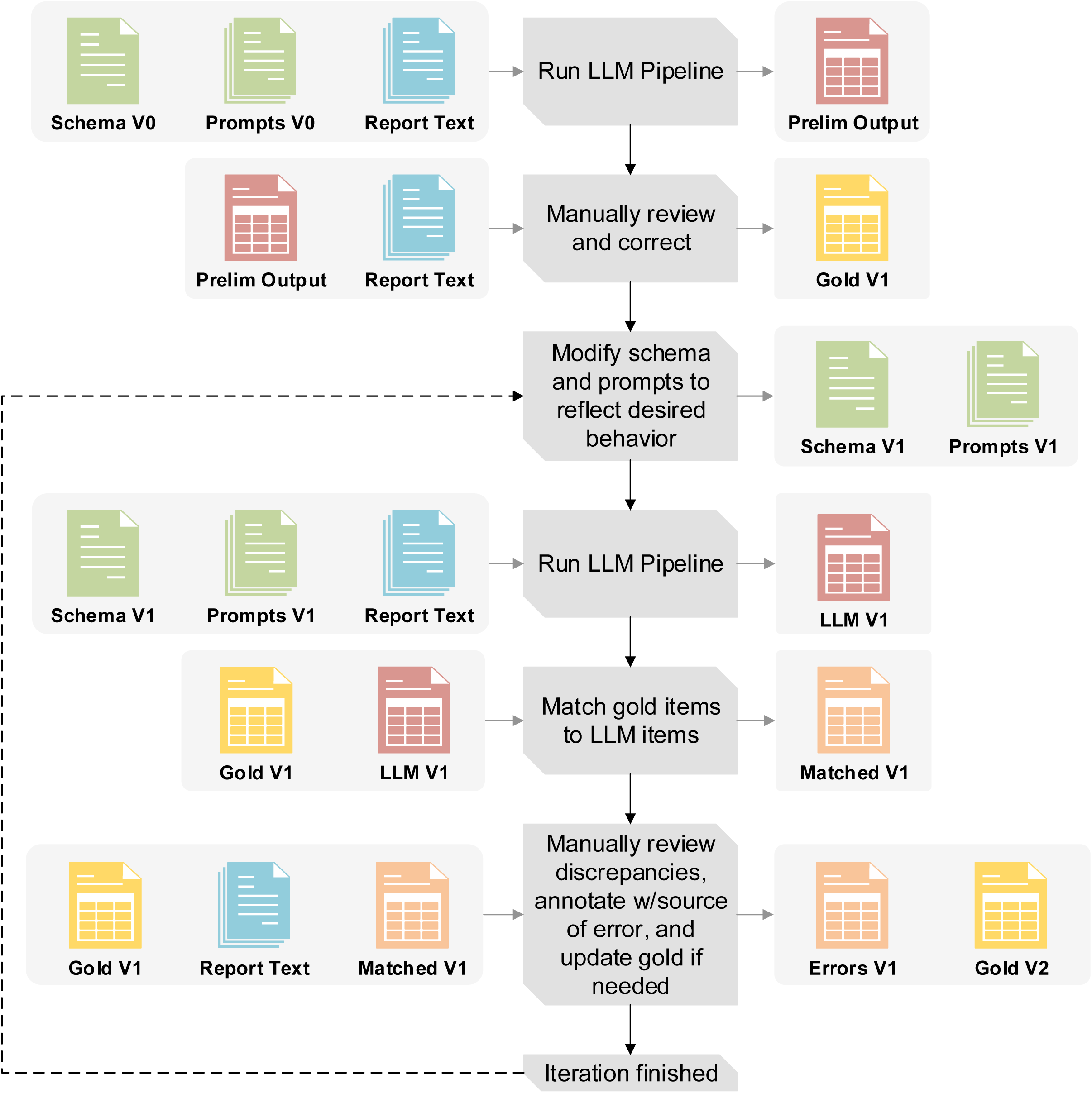
Overview of the iterative pipeline improvement and gold-standard set creation process. After completing each iteration, the schema, prompts, LLM outputs, and gold-standard are incrementally versioned e.g. V1, V2, V3, etc. The gold-standard set was structured in the exact format as the table that held the LLM outputs, with columns for report ID, specimen (and block when applicable) name, item name e.g histology, and item label e.g. clear cell RCC, so that items could be programmatically matched for review.

A data scientist (5 years clinical oncology experience) and a statistician (13 years kidney cancer research experience) reviewed annotations and discrepancies with all uncertainties or ambiguities deferred to a board-certified pathologist specializing in genitourinary pathology (23 years’ experience). All development used GPT-4o (2024-05-13, temperature 0) via a secure HIPAA compliant Azure API.^50^

### Creating an Error Ontology

A structured error ontology was developed to provide a framework for classifying the source, severity, and context of discrepancies between the LLM outputs and the gold-standard. The ontology comprises three sources of discrepancy: LLM, manual annotations (errors introduced to the gold-standard set by incorrect or insufficient annotation in a prior step), and what we term ‘schema issues’. Schema issues represent instances where the LLM and gold-standard were discordant, yet both appeared to have adhered to the provided instructions. In these cases, the instructions themselves were found to be insufficient or ambiguous. Both LLM and manual annotation discrepancies were further subclassified as of ‘major’ or ‘minor’ severity based on their potential impact on clinical interpretation or downstream analysis.

To provide finer details on the issues we encountered, we documented the contexts in which discrepancies arose. A flow chart for defining and documenting discrepancies as well as a brief introduction to the error context is provided in Figure 3. Detailed examples for a subset of contexts are given in Table 1, with the remainder plus additional examples in STable1. For each context, we first provide two potential labels for an entity arising from insufficient instructions in the given context. This is followed by our addressing methodology, and further examples of LLM or annotation error severity in similar contexts (provided that we found our instructions to be sufficient).

**Figure 3:**
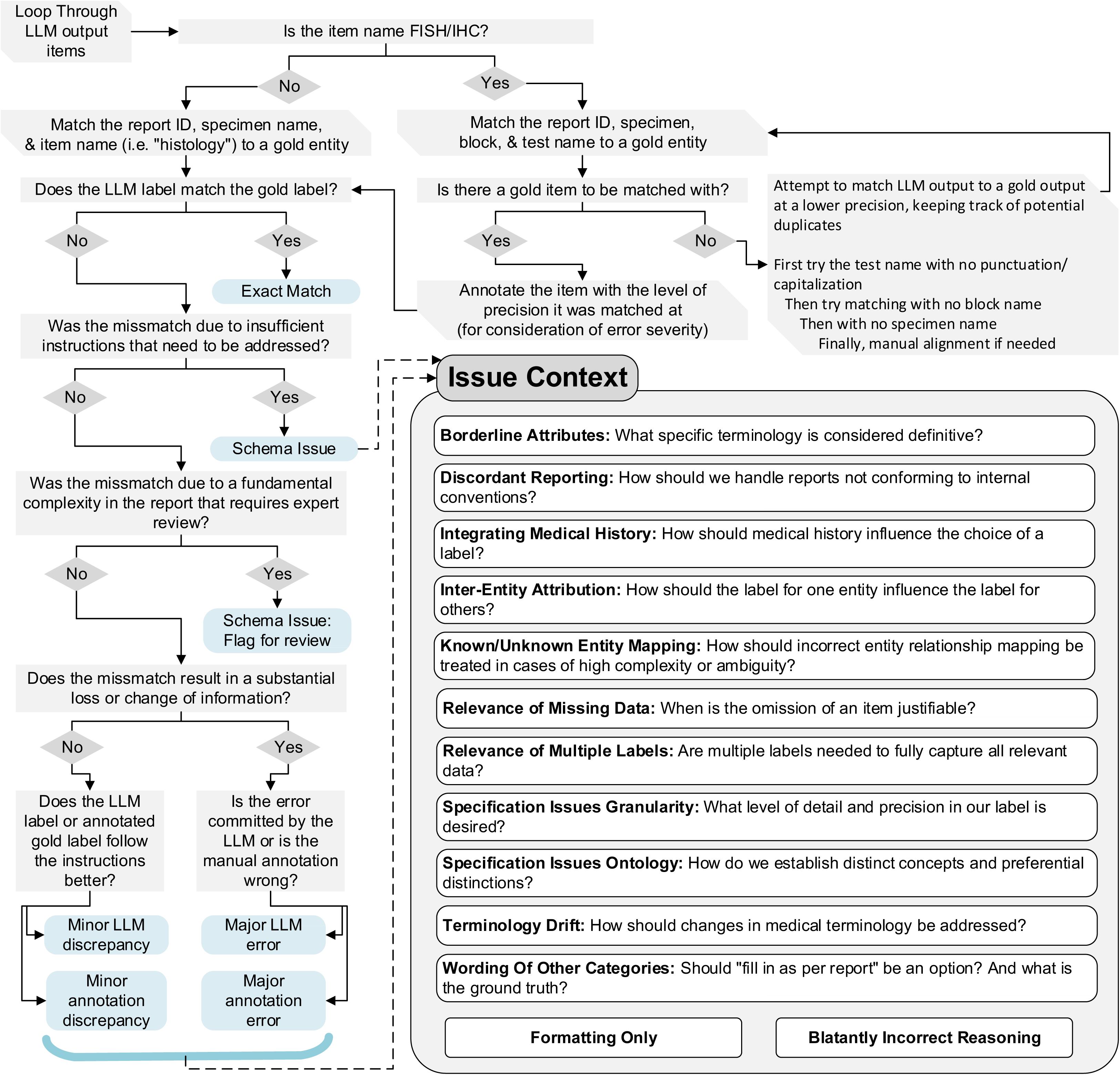
Flow chart for documenting discrepancy source and severity for an iteration. Issue contexts are introduced as questions needing to be asked about both workflow requirements and how certain kinds of deviations from instructions might need to be addressed. For the final two contexts: “Formatting only” refers to discrepancies that are purely due to standardized spellings/punctuation (BAP-1 vs BAP1), while “Blatantly Incorrect Reasoning” refers to errors not arising from any given nuanced context (e.g. hallucinating a test result not present in the report text)

**Table 1:** Issue Context Examples and Corrective Actions.

| Table 1: Issue Context Examples and Corrective Actions |  |  |
| --- | --- | --- |
| 1.1 Mixed Known/Unknown Entity Mapping |  |  |
| Report Text† | Review of outside slides<br>A. Skin, abdomen<br>- Metastatic carcinoma, IHC profile suggestive of renal primary<br>B. Skin, upper back<br>- Metastatic carcinoma, IHC profile suggestive of renal primary<br>...<br>IHC slides are positive for CK7, ... IHC stains were performed on block A2 and showed the following reactivity: PAX8 * Positive |  |
| Discordant Labels | X_block_X0_IHC_CK7: Positive<br>A_block_A2_IHC_PAX-8: Positive | A_block_A0_IHC_CK7: Positive<br>B_block_B0_IHC_CK7: Positive<br>A_block_A2_IHC_PAX-8: Positive |
| Context | - The initial schema instructed the use of specimen “X” as a stand-in when it is not clear which specimen was used for a test.<br>- In cases with multiple specimens of identical histology, for IHC tests lacking a specified specimen the LLM would continue to provide a duplicate set of results for all specimens. |  |
| Addressing Action | - A brief description of this situation along with a properly constructed output was added to the IHC/FISH segmentation II and standardization prompt. This new example provided additional reinforcement to maintain using X when specimen/block is not specified and the provided names only for the tests for which specimen/block correspondence is explicit. |  |
| Continued Error Severity Examples | - Major: If the duplicated set of results was returned for both A & B but B was benign tissue.<br>- Minor: Continued duplicated results, but only in the context of both specimens containing identical histology. |  |
| 1.2 Relevance of Multiple Labels |  |  |
| Report Text | A. Right kidney and adrenal gland, radical nephrectomy:<br>- Renal cell carcinoma, clear-cell type<br>- Adrenal gland, negative for malignancy |  |
| Discordant Labels | A_anatomical-site: Kidney, right;<br>Adrenal gland | A_anatomical-site: Kidney, right |
| Context | - The original instructions required listing all anatomical sites in the specimen, as some specimens have multiple anatomical sites.<br>- In the above report, the adrenal gland and kidney are anatomical sites in the same subpart- however only the kidney is positive for RCC.<br>- Ambiguity arose over whether to include both sites in the label for such contexts. |  |
| Addressing Action | - It was decided that for our purposes, we wanted the “anatomical site” field to continue to capture the primary organs/tissues removed for a specimen with no carve outs for histology. As such, in this case we would rely on the diagnosis and histology fields to guide our understanding that this was NOT a case of adrenal metastasis. |  |
| Continued Error Severity Examples | - Major: An anatomical site of only “Adrenal gland”, omitting the more important site.<br>- Minor: An anatomical site of only “Right kidney”. Although the adrenal gland is missing, because it is only benign tissue and not an RCC metastasis, its omission does not substantially affect planned downstream analysis. |  |
| (Intentionally blank space for table continuity) |  |  |

| 1.3 Specification Issue- Granularity |  |  |
| --- | --- | --- |
| Report Text | IHC performed on A2. Tumor cells are diffusely positive for CAIX in a membranous pattern |  |
| Discordant Labels | A_block_A2_IHC_CAIX: Positive, diffuse membranous | A_block_A2_IHC_CAIX: Positive, diffuse |
| Context | - In our original schema, we attempted to provide a list of all possible IHC results to choose from.<br>- After review we found this to be entirely impractical as the space of possible test results became enormous.<br>- We needed to precisely define the granularity of test results that we were interested in. |  |
| Addressing Action | - We shifted to a more modular schema comprising four dimensions—status, intensity, extent, and pattern—each with its own controlled vocabulary (see Figure 1A for an example).<br>- Under this new approach, the LLM is instructed to sequentially append any applicable modifiers (intensity, then extent, then pattern) to the primary status label, omitting those not present. |  |
| Continued Error Severity Examples | - Major: Returning only “Positive” as in RCC, we are very interested in detailed CAIX staining patterns.<br>- Minor: If in the example report CAIX had additional descriptors/modifiers that we are not interested in, thus are not in the schema, and are then returned by the LLM. These additional modifiers would not be factually incorrect, but would be beyond the standardized level of detail that we desire. |  |
| 1.4 Specification Issues- Ontology |  |  |
| Report Text | C. Peripancreatic mass, excision:<br>- Metastatic renal cell carcinoma, clear cell type |  |
| Discordant Labels | C_anatomical-site: Other-peripancreatic mass | C_anatomical-site: Pancreas |
| Context | - Because the mass is described as being peripancreatic, is it precise to label the site as pancreas?<br>- Additionally, in the context of metastasis, the histology of a tissue specimen should not be mistaken for its anatomical site |  |
| Addressing Action | - Added to anatomical site standardization instructions: “Analyze whether there are any position or direction terms that are relevant, for example a 'peripancreatic mass' would not be captured as 'Pancreas' as this refers to a mass in the tissue surrounding the pancreas. ... renal cell carcinoma that has metastasized to the left lung would ONLY have the anatomical site 'Lung, left' if the specimen ONLY contains lung tissue.” |  |
| Continued Error Severity Examples | - Major: Continued use of the label “pancreas” would be considered major as we have now instructed that the anatomical site must be consistent with the originating tissue.<br>- Minor: In some cases, continued usage of the “Other” label vs a specific provided label can be justified as a minor error if the site listed in the text does not cleanly map to labels in the schema. For example, an “intradural tumor” develops within the spinal cord, thus does not cleanly map to our schema label of “Spine, vertebral column” as this has a connotation of a tumor developing in bone tissue- although for our purpose we find this mapping acceptable. |  |
| (Intentionally blank space for table continuity) |  |  |

| 1.5 Integrating Medical History |  |  |
| --- | --- | --- |
| Report Text† | A. Soft tissue mass, parasplenic<br>- Poorly differentiated carcinoma, consistent with known renal cell carcinoma<br>Note: Prior history of papillary renal cell carcinoma is noted. |  |
| Discordant Labels | A_histology: Papillary renal cell carcinoma | A_histology: Poorly differentiated carcinoma |
| Context | - Should we label this specimen as papillary RCC inferring from the medical history, or only use the current report histology (poorly differentiated carcinoma)?<br>- The goal is to avoid automatically applying historical findings unless they are truly consistent with current specimen details. |  |
| Addressing Action | - Added to histology standardization instructions: “If a specimen is consistent or compatible with a known histology you may use that histology as part of your choice of a label, but ensure that the histology you choose is still applicable to the current specimen.” |  |
| Continued Error Severity Examples | - Major: If the report were to instead lack the “consistent with known renal cell carcinoma” modifier, then the histology “Papillary RCC” would be a major error as it would be reporting medical history alone.<br>- Minor: Labeling the specimen “RCC, no subtype specified” instead of “papillary RCC,” even though the text leans toward papillary (note the specimen is only consistent with *renal cell carcinoma*- no subtype specified). While not optimal, it does not fundamentally misclassify the specimen. |  |
| † Note that report text details and exact wording have been modified for brevity and to further enhance anonymity. |  |  |

### Stopping Criteria

Refinement concluded upon reaching zero major manual annotation errors, a near-zero rate of minor annotation discrepancies, a major LLM error rate near or below 1%, and an elimination of most schema errors, except those arising from complex cases deemed requiring human review. Upon reaching these criteria, we retrospectively reviewed the error documentation for each iteration to ensure consistency in our categorization (e.g., refining initial broad specification issues into more granular categories).

### Assessing LLM Interoperability

LLM backbone interoperability was assessed by comparing GPT-4o 2024-05-13, Llama 3.3 70B Instruct,^51^ and Qwen2.5 72B Instruct^52^ (running locally) outputs to the final gold-standard: First by exact match using the full specimen, block and test name when applicable, then by using an additional “fuzzy match” for IHC/FISH items that ignored punctuation and capitalization in the test name and did not require the specimen or tissue block identifiers to match the gold-standard. Exact match inter-model agreement between GPT-4o and the two open weight models was also calculated.

### Internal Application & Validation

Our finalized pipeline was run using GPT-4o 2024-08-06 on the free text portion (final diagnosis, ancillary studies, comments, addendums) of 3,520 internal pathology reports containing evidence of renal tumors spanning April 2019-April 2024; see SFigure 1 for details on inclusion criteria. Of these reports, 2,297 utilized additional discrete EMR fields corresponding to CAP kidney resection/biopsy and internal metastatic RCC pathology templates; This structured data could be pulled separately from the report text and was used for cross-referencing LLM output regarding metastatic RCC status and the presence or absence of six kidney tumor subtypes: clear cell RCC, chromophobe RCC, papillary RCC, clear cell papillary renal cell tumor (CCPRCT), TFE3-rearranged RCC, and TFEB-altered RCC. Discrepancies were manually reviewed using the free-text report as ground truth. Notably, the LLM needed to infer updated TFE3/TFEB subtypes from older ‘MiT family translocation RCC’ terms present in some structured data (per CAP Kidney 4.1.0.0).^43,44^

To provide a baseline for histology extraction and mapping to updated terminology, we developed a custom rule-based regular expression (regex) tool targeting predictable structural and lexical patterns commonly observed in the ‘Final Diagnosis’ section of reports, where primary histology is typically stated. This represented a pragmatic, non-machine learning approach using domain heuristics for comparison.

To attempt scalable validation of LLM extracted histology and IHC/FISH results across all 3,520 reports, including those with no available templated data, we selected all extracted subparts/specimens with a single histology of the above six for which IHC/FISH results were also extracted. We then assessed consistency of the histological subtype with the expected IHC/FISH pattern for 5 common markers used to differentiate RCC subtypes; CA-IX, CD117, Racemase, TFE3, and TFEB.^53^ Unexpected findings were subject to manual review of the report text.

### Assessing Clinical Domain Interoperability

Adaptability to different clinical domains was evaluated using The Cancer Genome Atlas (TCGA)^54–56^ Breast Invasive Carcinoma (BRCA)^57^ and Prostate Adenocarcinoma (PRAD)^58^ pathology reports that had undergone scanned PDF to text optical character recognition (OCR) processing and had corresponding tabular clinical data available.^59^ For BRCA reports, we extracted results for HER2 (both FISH and IHC separately), progesterone receptor (PR), and estrogen receptor (ER), utilizing only modified IHC/FISH schema labels and instructions. We restricted the reports to those containing the words “immunohistochemistry” and “HER2” to ensure IHC results were present in the OCR processed text.

PRAD reports were processed to extract Gleason Scores using the ‘feature report’ flow with only modifications to the schema instructions and labels. To gauge generalization, prompt templates were not modified and improvements were performed on only the schema, with iterations concluding once the schema adequately covered essential domain concepts and terminology. All external validation was done using GPT-4o (2024-08-06 via Azure, temperature of 0).

## Results

### Workflow Refinement & Gold-Standard Set

Following six iterative refinement cycles, our pipeline achieved strong alignment with the gold-standard annotations (1,413 total entities: 152 diagnoses, 651 specimen-level labels, 610 IHC/FISH results). The final iteration yielded a major LLM error rate of 0.99% (14/1,413) with no major annotation errors identified; Figure 4A. Schema issues necessitating more than flagging for review were also eliminated; Figure 4B, SFigures 2-4. STable 2 details distinct iteration updates with respect to fluctuating error prevalence.

**Figure 4:**
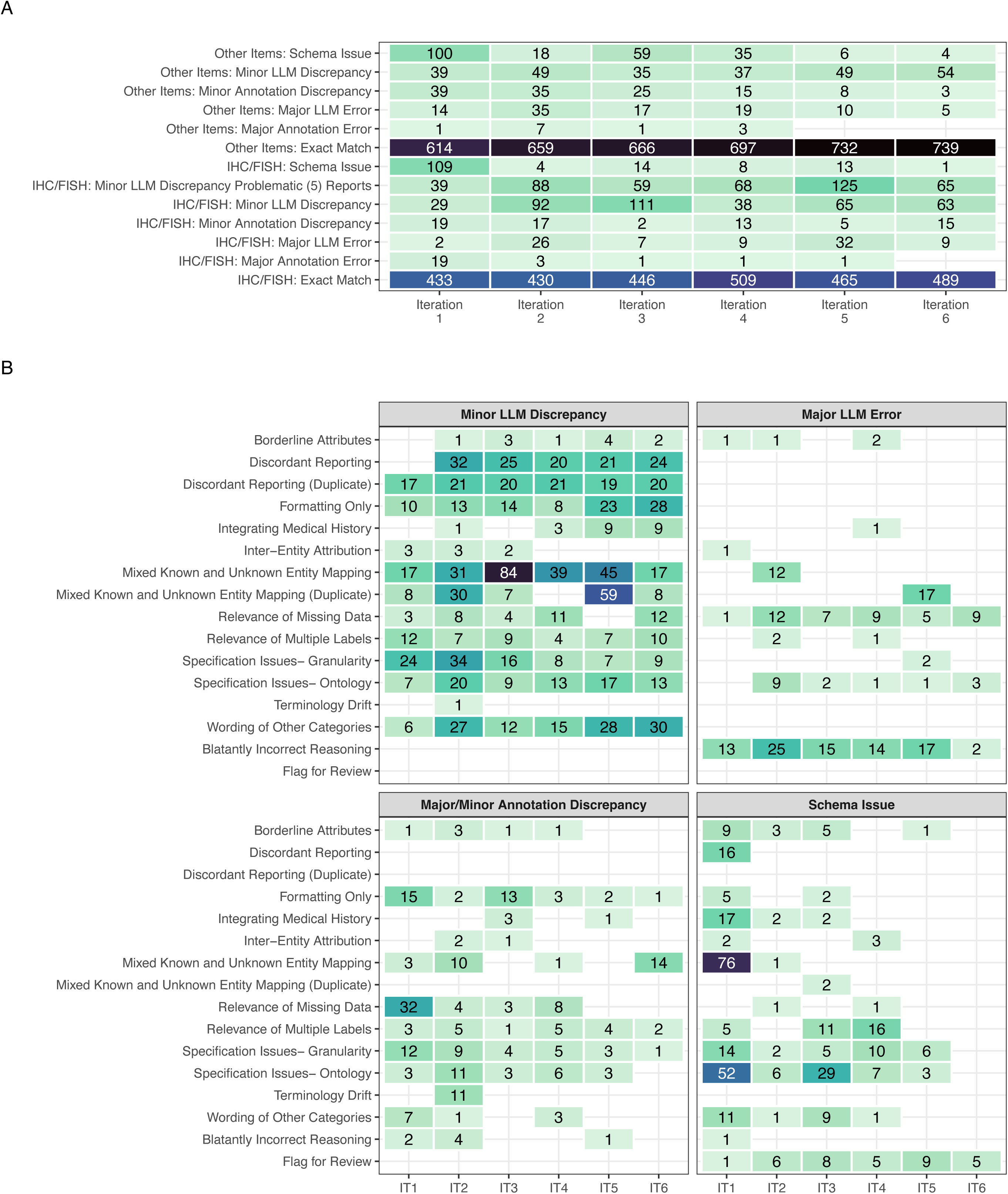
**(A)** Error/discrepancy source, severity, and entity type across iterations. Counts of 0 are left blank. Column totals are not equal across all iterations due to duplicate IHC/FISH entities and variations in missingness **(B)** Error/discrepancy contexts by source and severity across iterations (IT). Counts of 0 are left blank. Due to the lower number of major annotation errors, they have been grouped with minor annotation discrepancies for ease of visualization. For all panels, the fill color scale is maintained with a maximum at 84 and minimum of 1.

### Error Ontology

This iterative refinement process, systematically guided by our error ontology, provided critical insights into the diverse challenges encountered when extracting complex information from pathology reports. These challenges primarily fell into categories related to inherent report complexities, difficulties in task specification, normalization hurdles, and the integration of medical nuance.

#### Report Complexities

First, certain inherent report characteristics consistently generated discrepancies. Five complex outside consultations accounted for disproportionately many minor IHC/FISH discrepancies; Figure 4A. These ‘problematic reports’ all contained a mix of IHC/FISH tests where the associated block or specimen was clearly identified for some tests but not others. In such instances, rather than indicating ambiguity, the LLM would often incorrectly duplicate results across all specimens with similar histology; Table 1.1. Additionally, outside consultations often contained discordant reporting conventions for specimen names. For example, a specimen designated as “B” by the outside institution, could be referred to internally as specimen “A”; STable 1.1. Despite adding more illustrative examples of correctly mapped output to the prompts, these challenges persisted; Figure 4A, B. We also observed that major errors in IHC/FISH extraction, primarily missing results, were more prevalent in reports containing a high volume of tests (>10), and ambiguity arose in defining the severity of missing results when tests were mentioned by name but potentially not performed; STable 1.2.

#### Specification Issues

Second, precisely defining the desired information scope and granularity (‘specification issues’) was a key refinement focus. This required clarifying relevant entities when multiple labels were possible (e.g., anatomical sites, Table 1.2) and optimizing the level of detail for IHC results. The latter involved shifting from an exhaustive list of potential test results to a structured vocabulary capturing the distinct dimensions of status, intensity, extent, and pattern, allowing more flexible yet standardized representation; Table 1.3. Additional instructions were needed to handle nested labels, i.e. one label is more *precisely* correct than another, and the preferred level of granularity for capturing anatomical sites; Stables 1.3-4. Ontological nuances also required explicit instructions; for example, clarifying that a “peripancreatic mass” should not be coded as pancreatic metastasis required guidance on interpreting directional terms; Table 1.4.^60^ We also addressed ontological overlap, i.e. multiple labels could be considered *fully* correct, by establishing prioritization rules. For example, defining the preferred primary status for BAP-1 when both “Positive” and “Intact” were reported, STable 1.5. Finally, in certain circumstances when individual specimens shared characteristics, we needed to clarify when the label for one entity can affect that of another (e.g. Specimen A and B both originate from a “nephrectomy”, but in the report text specimen A’s procedure is referred to *only* as a “resection”; STable 1.6.

#### Normalization Difficulties

Third, normalizing terms and handling free-text entries remained challenging. While necessary to avoid information loss,^31^ the inclusion of “Other-<as per report>” categories consistently generated minor discrepancies due to the difficulty of achieving verbatim matches between the LLM output and gold-standard; STable 1.7.

Specific IHC/FISH term normalization, like standardizing ‘diffusely’ to ‘diffuse’, also proved problematic. This single normalization challenge accounted for over half of the remaining formatting-only discrepancies in the final iteration (15/28 errors) despite the target term ‘diffuse’ being relatively infrequent overall (46/610 IHC/FISH annotations). Investigating potential causes, we examined the GPT-4o tokenizer’s byte pair encoding (BPE) behavior. We found ‘diffusely’ tokenization varied with the preceding character: splitting into three tokens (’diff’, ‘us’, ‘ely’) after a newline but two (’diffus’, ‘ely’) after a space. While BPE segmentation is complex, we hypothesize this differential sub-word tokenization may contribute to the inconsistent application of this specific normalization rule. In contrast to these difficulties, the workflow proved highly adept at normalizing historical terminology to updated terms; Figure 4B, STable 1.8.

#### Medical Nuance

Finally, difficulties in integrating medical history and nuance required clinical domain expertise and corresponding adjustments. For example, the pathologist on our team clarified that in pathology reporting the terms ‘consistent with’ or ‘compatible with’ often carry more conclusive meaning than in general parlance, leading us to adjust instructions regarding the level certainty provided by these terms; STable 1.9.^61^ Similarly, interpreting complex distinctions, such as delineating local vs. distant lymph node metastases and assessing the relevance of medical history, necessitated providing detailed clinical context within the prompts; Table 1.5, STable 1.10-11.

### Assessing LLM Interoperability

The workflow proved variably flexible across LLM backbones. Compared to the gold-standard, exact match accuracies were 84.1% for GPT-4o (2024-08-06), 78.1% for Qwen-2.5 72B Instruct, and 70.1% for Llama 3.3 70B. Applying fuzzy matching for IHC/FISH items (ignoring test name punctuation/capitalization and relaxing specimen/block mapping criteria) improved these respective accuracies to 90.0%, 86.1%, and 82.0%. See STable 3.1 for exact counts and STable 3.2 for concrete examples of matching criteria. Exact match inter-model agreement between GPT-4o and the open-weight models was high (84.8% with Qwen-2.5, 82.2% with Llama 3.3). These results suggest the core prompt and schema logic is mostly transferable, though model choice impacts performance.

### Internal Application & Validation

Applying the finalized pipeline (using GPT-4o) to 2,297 internal kidney tumor reports with available structured EMR data for validation yielded high performance: a macro-averaged F1 score of 0.99 for identifying six key subtypes and an F1 of 0.97 for metastatic RCC detection; Table 2.1. Demonstrating clinical utility, the pipeline correctly identified necessary updates or corrections to this pre-existing structured data in 27 instances (e.g., reflecting addended results or more current terminology), all subsequently confirmed by a pathologist (Comprehensive details on discrepancies included in STable 4).

**Table 2.1:**
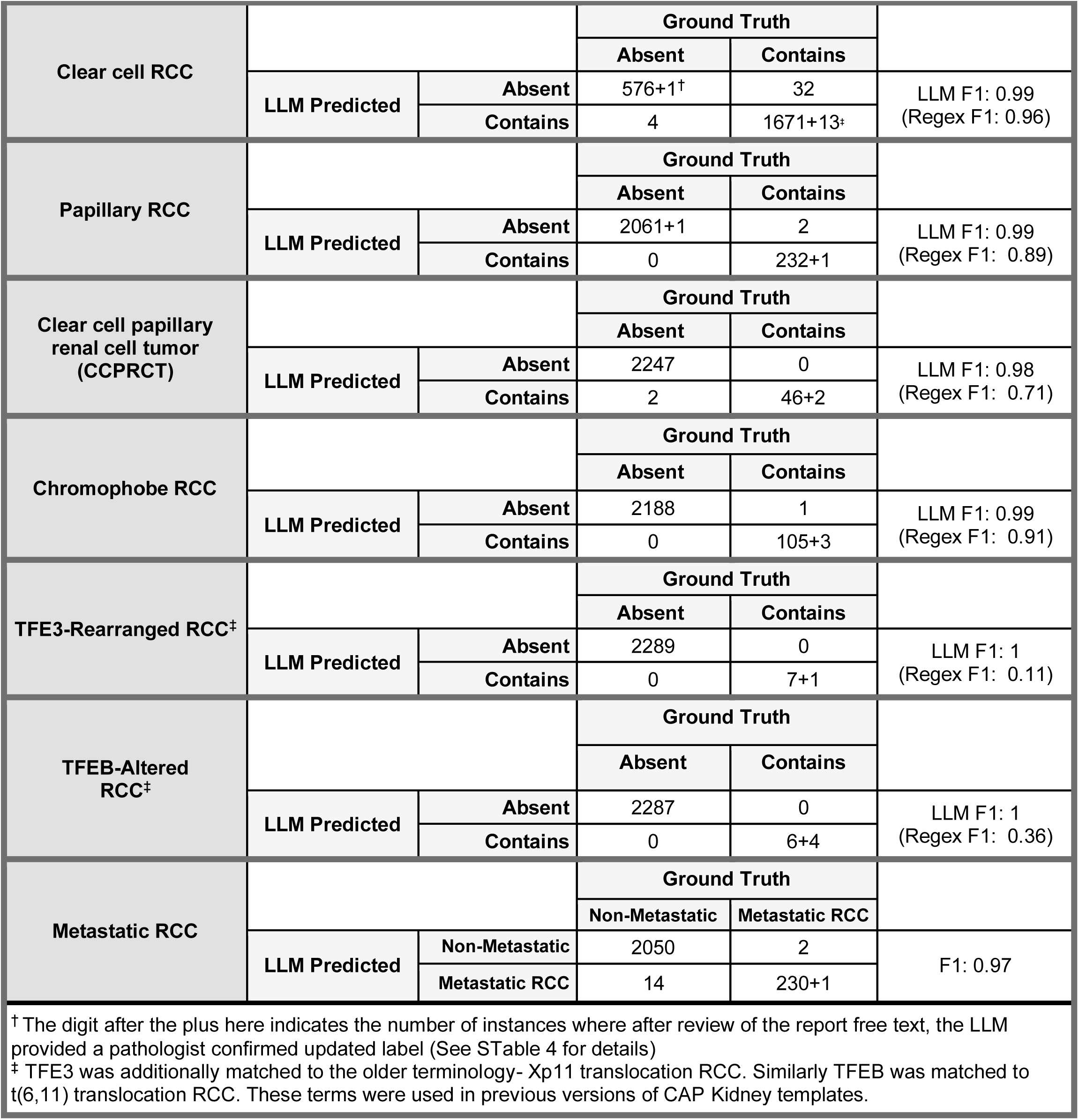
Consistency Between Preexisting Data and Extracted Histology and Diagnosis of Metastatic RCC.

A review of discrepancies for histological subtype showed continued difficulty with integrating medical history; most clear cell false negatives (28/32), for instance, resulted from incomplete use of a patient’s prior history of this subtype; STable 4. Lower performance in detecting metastatic RCC was primarily attributable to false positives due to misinterpreting medical history (n=6), local tumor extension (n=5), and differentiation of regional vs distant lymph nodes (n=3). Such errors often occurred in complex or ambiguous reports, underscoring the need for mechanisms to flag these cases for human review.

#### Comparison to Regex Baseline

The LLM pipeline demonstrated distinct advantages over the custom rule-based regex tool, particularly for less common subtypes or complex reports. The regex tool performed reasonably well for common subtypes with fewer historical variations in terminology (F1 scores: 0.96 Clear Cell, 0.91 Chromophobe, 0.89 Papillary RCC; Table 2.1). However, its performance degraded substantially when encountering wider terminology variation, historical naming conventions, or results reported primarily in comments or addendums – situations common for rarer subtypes. This resulted in much lower F1 scores for CCPRCT (0.71), TFE3-rearranged RCC (0.11), and TFEB-altered RCC (0.35), whereas the LLM pipeline maintained high accuracy and precision. Full regex results are available in STable 5.

#### Gauging Internal Consistency

To assess the reliability of extractions across the full set of 3,520 reports (including those without structured data for comparison), we examined internal consistency between extracted histology and IHC results. Out of all available reports, 2,464 subparts/specimens were identified as containing any of the histologies of interest. Of these, 1,906 were identified to have only a single histology and corresponding IHC results for the same subpart; SFigure 1. The pipeline showed a high degree of consistency, for example, 87/87 CD117 tests on specimens with chromophobe RCC were positive, and accurate extraction of the CA-IX ‘cup-like’ expected staining pattern for CCPRCT was demonstrated; Table 2.2. The two ‘box-like’ results found for CCPRCT corresponded to two tumors in a single report, wherein the LLM was consistent with the report text. The case was subsequently reviewed and found to have a ‘cup-like’ pattern and a correction was issued; STable 6.

**Table 2.2:**
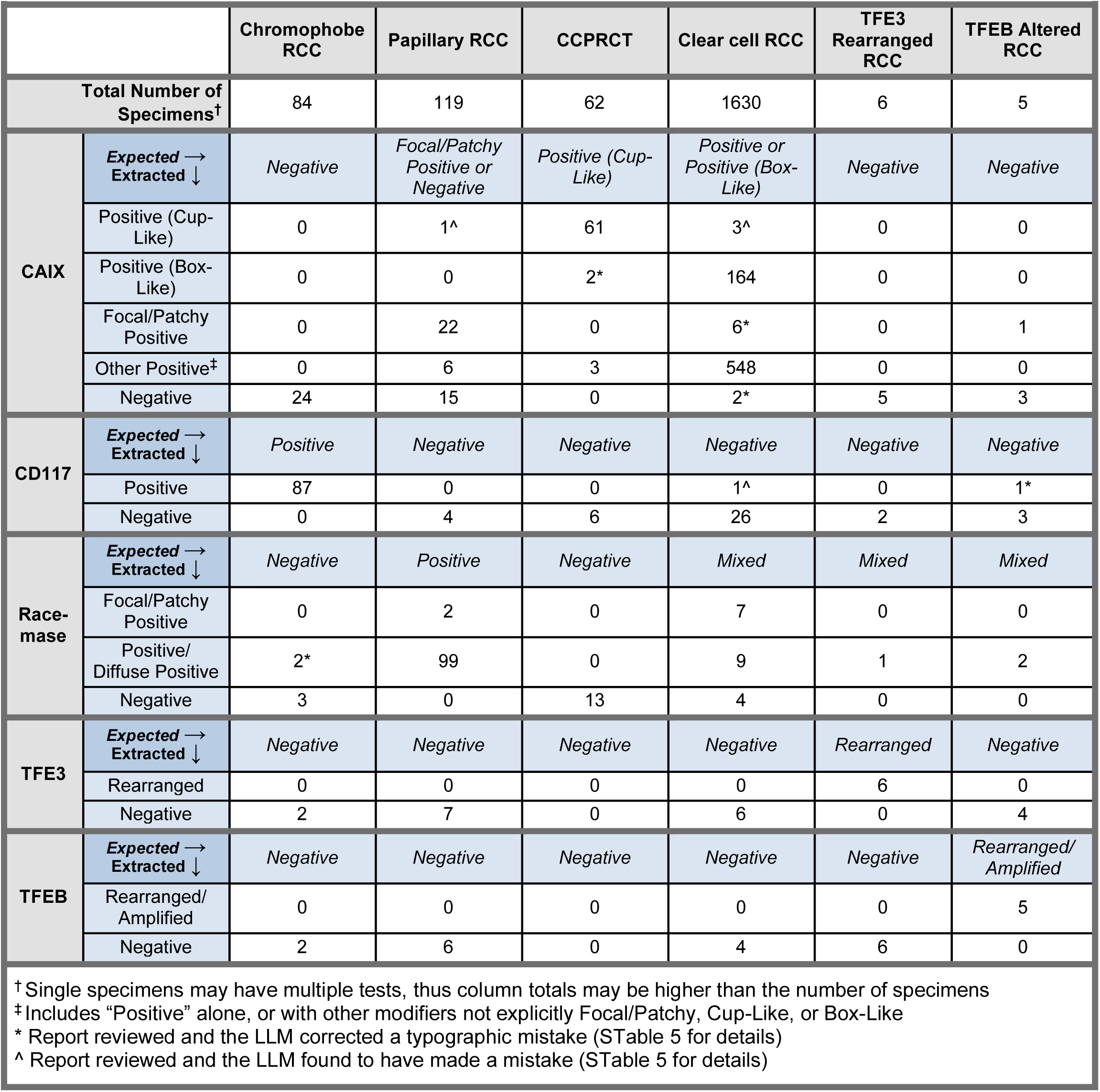
Consistency Between Extracted Histology and IHC/FISH Results.

### Assessing Clinical Domain Interoperability

Portability was assessed using publicly available TCGA data. Of the 757 available TCGA BRCA reports, 53 contained the words “immunohistochemistry” and “HER2” in the OCR text. Three schema only iterations were required to incorporate domain-specific terminology and reporting conventions for extracting HER2 (IHC and FISH), ER, and PR status. The pipeline (GPT-4o 2024-08-06) subsequently achieved 89% agreement with the curated tabular data; STable 7. Manual review of discrepancies suggested many were not LLM errors: nine cases involved results present in the curated data but absent in the scanned PDF, potentially indicating an alternate source for the curated result. These nine specific data points, where required information was not in the source text, were excluded from agreement calculations. Another result that first appeared to be an LLM false positive was subsequently found to reflect known clinical ambiguity regarding the classification of ER low-positive results (1-9% staining).^62^

Of the 333 available TCGA PRAD reports 253 had corresponding Gleason Scores in the structured clinical data and available OCR text. For this task, 98% agreement was achieved on the first run; STable 8. The difference in convergence speed (one run for PRAD vs. three for BRCA) highlights the impact of task and report complexity: extracting multiple, detailed IHC results from longer, more variable BRCA reports (average 9,250 characters) proved more challenging than extracting a single report-level Gleason Score from shorter PRAD reports (average 3,440 characters).

## Discussion

This study demonstrates that high accuracy in automated pathology report information extraction with large language models (LLMs) is possible but hinges on careful task definition and refinement. Although our pipeline yielded strong performance—for instance, a macro-averaged F1 score of 0.99 on identifying important RCC histological subtypes and adaptability to new entities such as Gleason Scores from prostate adenocarcinoma reports—our experience underscores that the process of defining information extraction goals may hold broader relevance than the performance metrics or workflow technicalities alone.

It became evident that the model’s success depended heavily on the clarity and depth of instructions. Consequently, a multidisciplinary team with domain expertise in NLP and LLMs, downstream statistical analysis, and clinical pathology became instrumental for success. Additional collaboration came from the LLM itself—particularly through review of the ‘reasoning’ output. We found that well-meaning instructions like “focus on the current specimen…, not past medical history” led to instances of ‘malicious compliance’ where the LLM followed instructions too literally, discarding important contextual information. Rectifying this required careful consideration of how instructions might be interpreted by the model, leading to increased specificity; Table 1.5. Underpinning this iterative process, the systematic application of our error ontology proved invaluable, not just for classifying discrepancies, but for prompting essential questions (as per Figure 3) and actively guiding the refinement of our extraction goals.

Cross-specialty collaboration was particularly vital for developing the error ontology. While developed with expert clinical input, we do acknowledge that our distinction between schema issues, major, and minor errors relied on contextual interpretation and our specific downstream use case. We thus caution against interpreting raw performance numbers in isolation. Instead, we advocate for a holistic interpretation considering the clinical significance of errors and their potential downstream impact, informed directly by stake-holding researchers.^63,64^ Interpretations could therefore differ between groups. For example, in broader kidney cancer research, deviations in the detailed staining pattern modifiers for CA-IX could be considered ‘major’ errors, as the distinction between ‘box-like’ vs ‘cup-like’ is decisive for RCC subtyping. However, if one were exclusively studying oncocytoma, where CA-IX is canonically negative,^65^ variations in the details of a ‘Positive’ result might sufficiently exclude this histology and be considered ‘minor’.

We note several limitations of our study and workflow. The iterative refinement process risked adding overly specific ‘one-off’ rules; we mitigated this by striving for generalizable instructions, using specific cases as examples rather than distinct rules; Table 1.4. Each refinement cycle also required highly detailed and time-intensive human review. One might ask whether an end-to-end manual annotation followed by traditional fine-tuning would have been easier. However, such an approach would have still demanded extensive manual review,^4^ offered no guarantee of easily handling the diverse and complex ‘error inducing’ contexts that we encountered, and might not have spurred the same level of insight into refining our information extraction goals. Moreover, generative LLM technology is evolving rapidly;^41^ our prompt-based pipeline is more adaptable to both changes in clinical entities and new model upgrades than static, fine-tuned architectures.

Furthermore, this approach currently produces only semi-structured data that in some instances requires further downstream normalization. While our research team possessed this domain-specific skillset, allowing us to prioritize granular detail over strict upfront standardization, this trade-off may not be acceptable for all teams and situations. Future work could leverage guided-decoding backends^37,38^ that can help constrain LLM generation to a predefined JSON schema. Such tools, along with increasing baseline AI performance and methods to flag problematic reports for human review, could further shift workflow efforts from managing formatting and data structuring requirements to well-informed task specification.

Limitations related to gold-standard development and validation also warrant mention. The standard was developed iteratively by the core team to refine complex task specifications, precluding formal inter-reviewer agreement metrics and limiting assessment of absolute annotation reliability. Our validation strategy—large-scale internal checks on core entities and external portability tests—aligned with the study’s focus on the refinement methodology and task specification challenges, rather than exhaustive benchmarking across every field against a static, held-out test set. As such, we posit that as LLMs are utilized to perform increasingly complex tasks, particularly those involving ambiguity and advancing medical knowledge, it may be useful to conceptualize ‘gold standard’ not merely as static ground truth, but as a dynamic reflection of evolving research goals and domain understanding.^40,63,64^

In summary, our LLM-based pipeline for pathology report information extraction highlights both strong performance metrics and the intricate processes required to achieve them. Our experience illustrates the importance of thoroughly understanding extraction intentions and goals, and how collaboration between domain experts—and even insights derived from the LLMs themselves—are crucial to this process. By documenting these complexities, we aim to provide a set of generalizable considerations that can inform future LLM-based clinical information extraction pipelines. As generative AI matures, flexible, human-in-the-loop strategies may prove essential to ensuring workflows remain grounded in real-world clinical objectives.

## Supporting information

SFigure 1

## Acknowledgments.

This work was supported by the NIH sponsored Kidney Cancer SPORE grant (P50CA196516) and endowment from Jan and Bob Pickens Distinguished Professorship in Medical Science and Brock Fund for Medical Science Chair in Pathology.

## Data Availability

The data used in this study contain patient identifiers and cannot be shared publicly due to privacy regulations and institutional policies.

The publicly available breast cancer reports and clinical data can be found at https://www.cbioportal.org/study/clinicalData?id=brca_tcga_pub2015

The publicly available prostate cancer reports and clinical data can be found at https://www.cbioportal.org/study/summary?id=prad_tcga_pub

And the optical character recognition processed reports are available at https://data.mendeley.com/datasets/hyg5xkznpx/1

## Code Availability

Code for implementing the workflow described in this paper is available at https://github.com/DavidHein96/prompts_to_table

## Author Contributions

DH: Conceptualization, Methodology, Data Collection, Data Analysis, Software, Visualization, Writing original draft, Writing review and editing

AC: Conceptualization, Data Collection, Data Analysis, Writing review and editing

MH: Software, Methodology, Writing review and editing

BX: Conceptualization, Data Collection, Writing review and editing

AJJ: Writing review and editing

JV: Data Collection, Clinical expertise, Writing review and editing

NR: Data Collection

AHS: Software

SC: Data collection, Software

LC: Conceptualization, Writing review and editing

JB: Writing review and editing, Project Administration & Supervision, Clinical expertise

AJ: Writing review and editing, Project Administration & Supervision

PK: Conceptualization, Data Collection, Writing review and editing, Project Administration & Supervision, Clinical expertise

## References

1. Li I, Pan J, Goldwasser J, et al. Neural Natural Language Processing for unstructured data in electronic health records: A review. Comput Sci Rev. 2022;46:100511. doi:10.1016/j.cosrev.2022.100511

2. Zozus MN, Pieper C, Johnson CM, et al. Factors Affecting Accuracy of Data Abstracted from Medical Records. Faragher EB, ed. PLOS ONE. 2015;10(10):e0138649. doi:10.1371/journal.pone.0138649

3. Brundin-Mather R, Soo A, Zuege DJ, et al. Secondary EMR data for quality improvement and research: A comparison of manual and electronic data collection from an integrated critical care electronic medical record system. J Crit Care. 2018;47:295–301. doi:10.1016/j.jcrc.2018.07.021

4. Sushil M, Kennedy VE, Mandair D, Miao BY, Zack T, Butte AJ. CORAL: Expert-Curated Oncology Reports to Advance Language Model Inference. NEJM AI. 2024;1(4). doi:10.1056/AIdbp2300110

5. Jee J, Fong C, Pichotta K, et al. Automated real-world data integration improves cancer outcome prediction. Nature. 2024;636(8043):728–736. doi:10.1038/s41586-024-08167-5

6. Sedlakova J, Daniore P, Horn Wintsch A, et al. Challenges and best practices for digital unstructured data enrichment in health research: A systematic narrative review. Sarmiento RF, ed. PLOS Digit Health. 2023;2(10):e0000347. doi:10.1371/journal.pdig.0000347

7. Xu H, Anderson K, Grann VR, Friedman C. Facilitating cancer research using natural language processing of pathology reports. Stud Health Technol Inform. 2004;107(Pt 1):565–572.

8. Hripcsak G, Friedman C, Alderson PO, DuMouchel W, Johnson SB, Clayton PD. Unlocking clinical data from narrative reports: a study of natural language processing. Ann Intern Med. 1995;122(9):681–688. doi:10.7326/0003-4819-122-9-199505010-00007

9. Alsentzer E, Murphy JR, Boag W, et al. Publicly Available Clinical BERT Embeddings. Published online 2019. doi:10.48550/ARXIV.1904.03323

10. Yang X, Chen A, PourNejatian N, et al. A large language model for electronic health records. Npj Digit Med. 2022;5(1):194. doi:10.1038/s41746-022-00742-2

11. Rasmy L, Xiang Y, Xie Z, Tao C, Zhi D. Med-BERT: pretrained contextualized embeddings on large-scale structured electronic health records for disease prediction. Npj Digit Med. 2021;4(1):86. doi:10.1038/s41746-021-00455-y

12. Lee J, Yoon W, Kim S, et al. BioBERT: a pre-trained biomedical language representation model for biomedical text mining. Wren J, ed. Bioinformatics. 2020;36(4):1234–1240. doi:10.1093/bioinformatics/btz682

13. Peng Y, Yan S, Lu Z. Transfer Learning in Biomedical Natural Language Processing: An Evaluation of BERT and ELMo on Ten Benchmarking Datasets. Published online 2019. doi:10.48550/ARXIV.1906.05474

14. Su P, Vijay-Shanker K. Investigation of improving the pre-training and fine-tuning of BERT model for biomedical relation extraction. BMC Bioinformatics. 2022;23(1):120. doi:10.1186/s12859-022-04642-w

15. Li Y, Wehbe RM, Ahmad FS, Wang H, Luo Y. A comparative study of pretrained language models for long clinical text. J Am Med Inform Assoc. 2023;30(2):340–347. doi:10.1093/jamia/ocac225

16. Brown TB, Mann B, Ryder N, et al. Language Models are Few-Shot Learners. Published online 2020. doi:10.48550/ARXIV.2005.14165

17. Liu H, Ning R, Teng Z, Liu J, Zhou Q, Zhang Y. Evaluating the Logical Reasoning Ability of ChatGPT and GPT-4. Published online 2023. doi:10.48550/ARXIV.2304.03439

18. Nori H, King N, McKinney SM, Carignan D, Horvitz E. Capabilities of GPT-4 on Medical Challenge Problems. Published online 2023. doi:10.48550/ARXIV.2303.13375

19. Singhal K, Azizi S, Tu T, et al. Large language models encode clinical knowledge. Nature. 2023;620(7972):172–180. doi:10.1038/s41586-023-06291-2

20. Agrawal M, Hegselmann S, Lang H, Kim Y, Sontag D. Large Language Models are Few-Shot Clinical Information Extractors. Published online 2022. doi:10.48550/ARXIV.2205.12689

21. Hu Y, Chen Q, Du J, et al. Improving large language models for clinical named entity recognition via prompt engineering. J Am Med Inform Assoc. 2024;31(9):1812–1820. doi:10.1093/jamia/ocad259

22. Sivarajkumar S, Kelley M, Samolyk-Mazzanti A, Visweswaran S, Wang Y. An Empirical Evaluation of Prompting Strategies for Large Language Models in Zero-Shot Clinical Natural Language Processing: Algorithm Development and Validation Study. JMIR Med Inform. 2024;12:e55318. doi:10.2196/55318

23. Peng C, Yang X, Chen A, et al. Generative large language models are all-purpose text analytics engines: text-to-text learning is all your need. J Am Med Inform Assoc. 2024;31(9):1892–1903. doi:10.1093/jamia/ocae078

24. Burford KG, Itzkowitz NG, Ortega AG, Teitler JO, Rundle AG. Use of Generative AI to Identify Helmet Status Among Patients With Micromobility-Related Injuries From Unstructured Clinical Notes. JAMA Netw Open. 2024;7(8):e2425981. doi:10.1001/jamanetworkopen.2024.25981

25. Hu D, Liu B, Zhu X, Lu X, Wu N. Zero-shot information extraction from radiological reports using ChatGPT. Int J Med Inf. 2024;183:105321. doi:10.1016/j.ijmedinf.2023.105321

26. Huang J, Yang DM, Rong R, et al. A critical assessment of using ChatGPT for extracting structured data from clinical notes. Npj Digit Med. 2024;7(1):106. doi:10.1038/s41746-024-01079-8

27. Johnson B, Bath T, Huang X, et al. Large language models for extracting histopathologic diagnoses from electronic health records. Published online November 28, 2024. doi:10.1101/2024.11.27.24318083

28. Le Guellec B, Lefèvre A, Geay C, et al. Performance of an Open-Source Large Language Model in Extracting Information from Free-Text Radiology Reports. Radiol Artif Intell. 2024;6(4):e230364. doi:10.1148/ryai.230364

29. Liu F, Li Z, Zhou H, et al. Large Language Models Are Poor Clinical Decision-Makers: A Comprehensive Benchmark. Published online April 25, 2024. doi:10.1101/2024.04.24.24306315

30. Omiye JA, Gui H, Rezaei SJ, Zou J, Daneshjou R. Large Language Models in Medicine: The Potentials and Pitfalls: A Narrative Review. Ann Intern Med. 2024;177(2):210–220. doi:10.7326/M23-2772

31. Sushil M, Zack T, Mandair D, et al. A comparative study of large language model-based zero-shot inference and task-specific supervised classification of breast cancer pathology reports. J Am Med Inform Assoc. 2024;31(10):2315–2327. doi:10.1093/jamia/ocae146

32. Wang LL, Otmakhova Y, DeYoung J, et al. Automated Metrics for Medical Multi-Document Summarization Disagree with Human Evaluations. Proc Conf Assoc Comput Linguist Meet. 2023;2023:9871–9889. doi:10.18653/v1/2023.acl-long.549

33. Wornow M, Xu Y, Thapa R, et al. The shaky foundations of large language models and foundation models for electronic health records. Npj Digit Med. 2023;6(1):135. doi:10.1038/s41746-023-00879-8

34. Tang L, Sun Z, Idnay B, et al. Evaluating large language models on medical evidence summarization. Npj Digit Med. 2023;6(1):158. doi:10.1038/s41746-023-00896-7

35. Reichenpfader D, Müller H, Denecke K. A scoping review of large language model based approaches for information extraction from radiology reports. Npj Digit Med. 2024;7(1):222. doi:10.1038/s41746-024-01219-0

36. Tian S, Jin Q, Yeganova L, et al. Opportunities and challenges for ChatGPT and large language models in biomedicine and health. Brief Bioinform. 2023;25(1):bbad493. doi:10.1093/bib/bbad493

37. Willard BT, Louf R. Efficient Guided Generation for Large Language Models. Published online 2023. doi:10.48550/ARXIV.2307.09702

38. Dong Y, Ruan CF, Cai Y, et al. XGrammar: Flexible and Efficient Structured Generation Engine for Large Language Models. Published online 2024. doi:10.48550/ARXIV.2411.15100

39. Fleming SL, Lozano A, Haberkorn WJ, et al. MedAlign: A Clinician-Generated Dataset for Instruction Following with Electronic Medical Records. Published online 2023. doi:10.48550/ARXIV.2308.14089

40. McIntosh TR, Susnjak T, Arachchilage N, Liu T, Watters P, Halgamuge MN. Inadequacies of Large Language Model Benchmarks in the Era of Generative Artificial Intelligence. Published online 2024. doi:10.48550/ARXIV.2402.09880

41. Zhong T, Liu Z, Pan Y, et al. Evaluation of OpenAI o1: Opportunities and Challenges of AGI. Published online 2024. doi:10.48550/ARXIV.2409.18486

42. Goel A, Gueta A, Gilon O, et al. LLMs Accelerate Annotation for Medical Information Extraction. Published online 2023. doi:10.48550/ARXIV.2312.02296

43. Murugan P. Protocol for the Examination of Resection Specimens from Patients with Renal Cell Carcinoma. Published online June 2024. https://documents.cap.org/protocols/Kidney_4.2.0.0.REL_CAPCP.pdf

44. Murugan P. Protocol for the Examination of Biopsy Specimens from Patients with Renal Cell Carcinoma. Published online June 2024. https://documents.cap.org/protocols/Kidney.Bx_4.2.0.0.REL_CAPCP.pdf

45. Prompt flow. https://github.com/microsoft/promptflow?tab=readme-ov-file

46. Wei J, Wang X, Schuurmans D, et al. Chain-of-Thought Prompting Elicits Reasoning in Large Language Models. Published online 2022. doi:10.48550/ARXIV.2201.11903

47. Yu Z, He L, Wu Z, Dai X, Chen J. Towards Better Chain-of-Thought Prompting Strategies: A Survey. Published online 2023. doi:10.48550/ARXIV.2310.04959

48. Chen Q, Qin L, Liu J, et al. Towards Reasoning Era: A Survey of Long Chain-of-Thought for Reasoning Large Language Models. Published online 2025. doi:10.48550/ARXIV.2503.09567

49. Yeo WJ, Satapathy R, Goh RSM, Cambria E. How Interpretable are Reasoning Explanations from Prompting Large Language Models? Published online 2024. doi:10.48550/ARXIV.2402.11863

50. OpenAI, Hurst A, Lerer A, et al. GPT-4o System Card. Published online 2024. doi:10.48550/ARXIV.2410.21276

51. Grattafiori A, Dubey A, Jauhri A, et al. The Llama 3 Herd of Models. Published online 2024. doi:10.48550/ARXIV.2407.21783

52. Yang A, Yang B, Hui B, et al. Qwen2 Technical Report. Published online 2024. doi:10.48550/ARXIV.2407.10671

53. Kim M, Joo JW, Lee SJ, Cho YA, Park CK, Cho NH. Comprehensive Immunoprofiles of Renal Cell Carcinoma Subtypes. Cancers. 2020;12(3):602. doi:10.3390/cancers12030602

54. De Bruijn I, Kundra R, Mastrogiacomo B, et al. Analysis and Visualization of Longitudinal Genomic and Clinical Data from the AACR Project GENIE Biopharma Collaborative in cBioPortal. Cancer Res. 2023;83(23):3861–3867. doi:10.1158/0008-5472.CAN-23-0816

55. Cerami E, Gao J, Dogrusoz U, et al. The cBio Cancer Genomics Portal: An Open Platform for Exploring Multidimensional Cancer Genomics Data. Cancer Discov. 2012;2(5):401–404. doi:10.1158/2159-8290.CD-12-0095

56. Gao J, Aksoy BA, Dogrusoz U, et al. Integrative Analysis of Complex Cancer Genomics and Clinical Profiles Using the cBioPortal. Sci Signal. 2013;6(269). doi:10.1126/scisignal.2004088

57. Ciriello G, Gatza ML, Beck AH, et al. Comprehensive Molecular Portraits of Invasive Lobular Breast Cancer. Cell. 2015;163(2):506–519. doi:10.1016/j.cell.2015.09.033

58. Abeshouse A, Ahn J, Akbani R, et al. The Molecular Taxonomy of Primary Prostate Cancer. Cell. 2015;163(4):1011–1025. doi:10.1016/j.cell.2015.10.025

59. Kefeli J, Tatonetti N. TCGA-Reports: A machine-readable pathology report resource for benchmarking text-based AI models. Patterns. 2024;5(3):100933. doi:10.1016/j.patter.2024.100933

60. Lavu H, Yeo CJ. Metastatic renal cell carcinoma to the pancreas. Gastroenterol Hepatol. 2011;7(10):699–700.

61. Oien KA, Dennis JL. Diagnostic work-up of carcinoma of unknown primary: from immunohistochemistry to molecular profiling. Ann Oncol. 2012;23:x271–x277. doi:10.1093/annonc/mds357

62. Makhlouf S, Althobiti M, Toss M, et al. The Clinical and Biological Significance of Estrogen Receptor-Low Positive Breast Cancer. Mod Pathol. 2023;36(10):100284. doi:10.1016/j.modpat.2023.100284

63. Shool S, Adimi S, Saboori Amleshi R, Bitaraf E, Golpira R, Tara M. A systematic review of large language model (LLM) evaluations in clinical medicine. BMC Med Inform Decis Mak. 2025;25(1):117. doi:10.1186/s12911-025-02954-4

64. Ho CN, Tian T, Ayers AT, et al. Qualitative metrics from the biomedical literature for evaluating large language models in clinical decision-making: a narrative review. BMC Med Inform Decis Mak. 2024;24(1):357. doi:10.1186/s12911-024-02757-z

65. Büscheck F, Fraune C, Simon R, et al. Aberrant expression of membranous carbonic anhydrase IX (CAIX) is associated with unfavorable disease course in papillary and clear cell renal cell carcinoma. Urol Oncol Semin Orig Investig. 2018;36(12):531.e19–531.e25. doi:10.1016/j.urolonc.2018.08.015

