## Supplemental Appendix for "Prompts to Table: Specification and Iterative Refinement for Clinical Information Extraction with Large Language Models"

### STable 1: Issue Context Examples and Corrective Actions Continued

#### 1.1 Discordant Reporting Practices

| | | |
|---|---|---|
| **Report Text** | Outside case for consultation (Original case number XX:SXXXXXX)<br>**A**. Lymph node; biopsy (XX:SXXXXXX, **B1**): IHC positive for CA-IX<br>**B**. Liver; biopsy (PL:S12345, **C1**): Benign tissue | |
| **Discordant Labels** | **A**_block_**A0**_IHC_CA-IX: Positive | **A**_block_**B1**_IHC_CA-IX: Positive |
| **Context** | - In this outside consultation, the outside specimen consists of a “B” specimen and a “C” specimen, but they are provided with internal names as “A” and “B”.<br>- The LLM sees “B1” and interprets this as a block name, however this specimen name is contradicted by the internal specimen name. | |
| **Addressing Action** | - Added to IHC/FISH segmentation II and standardization prompts: Instructions and examples on always using the internal name if provided, and using specimen “X” if no internal names are provided. | |
| **Continued Error Severity Examples** | **Major:** Attributing this result to specimen B, as B is benign tissue<br>**Minor:** If in the example report, both specimen A and B were nearly identical tissue types and histologies, depending on the context (say internal names were not provided at all or it is unclear which specimen was used), misattribution of block and specimen names may be considered a minor error. | |

#### 1.2 Relevance of Missing Data

| | | |
|---|---|---|
| **Report Text** | Low-grade oncocytic tumor (LOT) category has emerged from such category in recent years. This tumor is characterized by diffuse CK7 positivity. I would recommend performing this immunohistochemical stain in the largest tumor to rule out the possibility of LOT. | |
| **Discordant Labels** | X_block_X0_IHC_CK7: Result not provided | *(No result)* |
| **Context** | - The initial schema and instructions to reviewers was to record all tests without results as having the status “Result not provided”.<br>- However, in some cases a test is only recommended, as is CK7 in the above report, thus there is not technically a “Result” to not provide. | |
| **Addressing Action** | - Added to IHC/FISH segmentation II prompt: “Is any text actually a reference for interpreting the outcome of a test? If so, this is not a test result and should not be included in the answer.” | |
| **Continued Error Severity Examples** | Major: Returning “CK7 Positive, diffuse”, misinterpreting the reference as a result.<br>Minor: Continued inclusion of this test, with a result such as “Result not provided” could be a minor error. Technically, there is no result to not provide, but for our purposes, it does not substantially change the information extracted from the report. | |

#### 1.3 Specification Issues Granularity II: Overlapping Labels II (Nested/subset labels)

| | | |
|---|---|---|
| **Report Text** | `A. Right kidney, resection. eosinophilic vacuolated tumor (EVT)` | |
| **Discordant Labels** | `A_histology: Other oncocytic tumors of the kidney` | `A_histology: Other- eosinophilic vacuolated tumor` |
| **Context** | - This specific histology is not found in the RCC CAP template but would technically fall under ‘Other oncocytic tumors of the kidney’ which *is* included in the template.<br>- In this case there is one more specific label that is correct, however a second more broad label is also correct. | |
| **Addressing Action** | - Added to the histology standardization instructions “When there is *truly no good match* for a specimen, return 'Other- <fill in as per report>' and fill in the specific histology as per the report” to promote using a specific label over the other category. | |
| **Continued Error Severity Examples** | **Major:** Returning “Benign tissue” if the LLM were to reason that since the reported histology is not in the provided list of RCC subtypes, it must in fact be a benign histology.<br>**Minor:** Continued label discrepancy due to use of the dedicated “Other oncocytic tumors…” label vs the “Other-<fill in the blank” could be considered minor if there is no loss in information. | |

#### 1.4 Specification Issues Granularity III: An Extra Example

| | | |
|---|---|---|
| **Report Text** | `A. Left atrial mass` | |
| **Discordant Labels** | `A_anatomical-site: Heart` | `A_anatomical-site: Other- left atrial mass` |
| **Context** | - In this situation we desire a certain level of specification to enable efficient downstream analysis.<br>- When do we want an entity to be labeled with a more general term from the standardized list vs use the ‘other’ category and be more specific? | |
| **Addressing Action** | - Added to the anatomical site standardization instructions: “Try to capture the site at the level of granularity of the provided list. For example, if the site is cerebellum in the text, return 'Brain', as the cerebellum is part of the brain … When capturing a site at a less detailed level of granularity, ensure that you explain your reasoning and reflect on how you came to the conclusion, think about how you would explain it to a fifth grader.” | |
| **Continued Error Severity Examples** | **Major:** An anatomical site outside of the heart<br>**Minor:** Returning “Heart, left atrium” would not conform to the level of granularity desired, but would not be technically incorrect. | |

##### 1.5 Specification Issues Ontology II: Overlapping Labels I (Two equally correct labels)

| | | |
|---|---|---|
| **Report Text** | `A. Right kidney, partial nephrectomy`<br>`IHC performed on block A1: BAP-1 Intact (Positive nuclear staining)` | |
| **Discordant Labels** | `A_block_A1_IHC_BAP-1: Intact` | `A_block_A1_IHC_BAP-1: Positive` |
| **Context** | - Many IHC tests are reported as either positive OR intact. However, the common RCC test BAP-1 uses a particular wording “Intact (Positive nuclear staining)”.<br>- As both Positive and Intact are “status” options in the IHC results, there was an inconsistency in which term was used. There are two distinct correct labels. | |
| **Addressing Action** | - Added to IHC/FISH standardization instructions: “the result 'Intact (Positive nuclear staining ...)' favor returning only the result 'Intact', no other qualifiers are needed. Only do this if BOTH the words Intact AND Positive are used together” | |
| **Continued Error Severity Examples** | **Major:** A test result outside of the overlapping labels- “Negative”<br>**Minor:** Continued use of the label “Positive” in this situation, as long as “Positive” is present in the report text. | |

##### 1.6 Inter-Entity Attribution

| | | |
|---|---|---|
| **Report Text** | `A. Kidney, left renal tissue, possible tumor, resection:`<br>`B. Kidney, left renal mass, partial nephrectomy` | |
| **Discordant Labels** | `A_procedure: Resection, not otherwise specified`<br>`B_procedure: Partial nephrectomy` | `A_procedure: Partial nephrectomy`<br>`B_procedure: Partial nephrectomy` |
| **Context** | - Specimen B explicitly states “partial nephrectomy,” while Specimen A only says “resection” despite both originating from the same kidney/procedure.<br>- Should we infer the procedure for A from B, or strictly use the text for each specimen independently?<br>- Inferring across specimens was initially commonly done by annotators | |
| **Addressing Action** | - Annotators instructed to strictly use the procedure as stated for each specimen. | |
| **Continued Error Severity Examples** | **Major:** A procedure of “Resection, not otherwise specified” for specimen B, resulting in an important loss of information.<br>**Minor:** Use of the label “Partial nephrectomy” for specimen A after schema updating. This is not technically incorrect but does not align with the instructions. | |

#### 1.7 Wording of “Other” Categories

<table>
<tr><td>Report Text</td><td colspan="2"><code>B. Liver, resection: -Metastatic carcinoma, consistent with Wilms tumor</code></td></tr>
<tr><td>Discordant Labels</td><td><code>diagnosis: Other- Metastatic Wilms tumor</code></td><td><code>diagnosis: Other- Wilms tumor with metastasis to liver</code></td></tr>
<tr><td>Context</td><td colspan="2">- Wilms tumor is an uncommon diagnosis and is not an RCC, and further, does not neatly fit into any other diagnosis options, thus requiring an “Other-<fill in the blank>” schema label.<br>- Variations in how “Other” is worded (e.g., “Metastatic Wilms tumor” vs. “Wilms tumor with liver metastasis”) can cause mismatches, even with case insensitivity.</td></tr>
<tr><td>Addressing Action</td><td colspan="2">- For the feature report and feature specimen template: Instruct both human reviewers and the LLM to closely follow the report’s exact phrasing when applying “Other-” e.g. “ filling in with the actual {{feature}} as stated in the report”.<br>- In the example provided, “Metastatic Wilms tumor” is favored as it more closely matches the exact wording in the report.</td></tr>
<tr><td>Continued Error Severity Examples</td><td colspan="2">Major: A diagnosis of “Other- metastatic liver carcinoma”, this wording of the other category is clinically incorrect.<br>Minor: Minor wording differences (e.g., “Other- Wilms tumor with metastasis to liver”) that do not alter the essential meaning.</td></tr>
</table>

#### 1.8 Terminology Drift

<table>
<tr><td>Report Text</td><td colspan="2"><code>A. Left kidney, ...  - Xp11 translocation renal cell carcinoma</code></td></tr>
<tr><td>Discordant Labels</td><td><code>A_histology: TFE3-rearranged renal cell carcinomas</code></td><td><code>A_histology: Xp11 translocation renal cell carcinoma</code></td></tr>
<tr><td>Context</td><td colspan="2">- Prior to June 2024, the CAP template included the histology Xp11 translocation RCC.<br>- As of June 2024, this was replaced with TFE3-rearranged RCC.</td></tr>
<tr><td>Addressing Action</td><td colspan="2">- Added to histology standardization instructions:  “For your reference, Xp11 translocation renal cell carcinoma is now referred to as TFE3-rearranged renal cell carcinoma” was added to the schema.</td></tr>
<tr><td>Continued Error Severity Examples</td><td colspan="2">Major: A histology of “TFEB-altered RCC”, an incorrect updated histology.<br>Minor: Continued use of the “Xp11” label would not be factually incorrect, but would not align with standardization goals.</td></tr>
</table>

##### 1.9 Borderline Attributes

| | | |
|---|---|---|
| **Report Text** | Per outside report, the tumor cells are positive for CAM 5.2, and negative for cathepsin K, Melan-A, HMB-45, CK7. The morphology of this high grade infiltrative RCC is not classic for clear cell subtype. We repeated CA-IX and BAP1 IHC at our institution. CA-IX showed a diffuse membranous staining in zonal areas and BAP1 nuclear expression was lost. Though the tumor does not show the classic vasculature of ccRCC and diffuse CA-IX expression, the mutation profile is compatible with clear cell subtype. | |
| **Discordant Labels** | A_histology: Clear cell renal cell carcinoma | A_histology: Renal cell carcinoma, unclassified (NOS) |
| **Context** | - Our original schema was strict about only using RCC subtypes when they were "definitively confirmed"<br>- However, pathologists often use phrases such as "compatible with" or "consistent with" that in day-to-day language, have a slightly lower degree of certainty than "confirmed". | |
| **Addressing Action** | - Added to histology segmentation instructions: "reflect in your reasoning on the strength of certainty of the pathologist … There may be an addendum to the report that will confirm or deny the potential histologies"<br>- Added to histology standardization instructions: "Terms like 'prior history noted' or 'suggestive of' may not be strong enough to conclusively diagnose a certain histology. Reflect in your reasoning on the strength of certainty of the pathologist." | |
| **Continued Error Severity Examples** | **Major:** After clarifying this wording in the schema, RCC NOS would be considered a major error.<br>**Minor:** Not shown in the example, but depending on the context and available IHC stains "suggestive of"- may or may not be strong enough to warrant labeling with a specific subtype and a mismatch could be considered minor. In such cases, the desired behavior would actually be to flag for human review. | |

##### 1.10 Medical Nuance: Regional vs Distant Metastasis

| | | |
|---|---|---|
| **Report Text** | A. Lymph node, left para-aortic, CT-guided needle biopsy:<br>- Metastatic carcinoma consistent with Chromophobe renal cell carcinoma<br>...<br>The patient has a prior pathology diagnosis on left radical nephrectomy of RCC. The morphology of this metastasis is similar to the nephrectomy specimen | |
| **Discordant Labels** | diagnosis: RCC with regional lymph node metastasis | diagnosis: Metastatic RCC |
| **Context** | - For our use case, we desire a differentiation between regional lymph node metastasis and distant metastasis<br>- Additionally, this case contains a metastasis to a lymph node that is regional, however RCC staging is done at time of diagnosis/cancer resection and considered a recurrence/metastasis post nephrectomy. | |
| **Addressing Action** | - Added to the diagnosis standardization instructions- "If there is confirmed RCC that has only spread to regional lymph nodes- hilar, Precaval, Interaortocaval, Paracaval, Retrocaval, Preaortic, Paraaortic, Retroaortic, retroperitoneal- diagnose as RCC with regional lymph node metastasis" | |
| **Continued Error Severity Examples** | **Major:** We are very interested in this distinction, so any incorrect label would be considered major.<br>**Minor:** NA | |

##### 1.11 Integrating Medical History II: References to Other Reports

| | | |
|---|---|---|
| **Report Text** | A. Right renal mass (radical nephrectomy): Clear cell renal cell carcinoma<br>AJCC 8th edition pathologic staging: pT3a N0<br>The patients prior biopsy (SU-123456) from liver metastasis was concurrently reviewed and the current case shows similar findings | |
| **Discordant Labels** | diagnosis: Malignant neoplasm of right kidney, except renal pelvis | diagnosis: Metastatic RCC |
| **Context** | - The current report only contains kidney tissue, but references a separate liver biopsy indicating metastatic RCC.<br>- Should the pipeline label the current diagnosis as metastatic RCC based on history, or strictly reflect findings from the present specimen?<br>- Conversely, if the report is benign but references a prior metastasis, do we label it metastatic? | |
| **Addressing Action** | - Typically we want the diagnosis for a report to reflect only the report findings.<br>- For our purposes, we maintain instructions to "focus only on the current report," especially to avoid false positives for metastatic disease. | |
| **Continued Error Severity Examples** | **Major:** Not shown, but labeling a benign kidney sample as metastatic RCC solely due to external references.<br>**Minor:** Not shown in the above example, but if a report were to contain a single specimen with clear cell, and a reference to another specimen in a different report with the same histology, the inclusion of both specimens in the output could be a minor error. Ideally, only specimens in the current report are returned, but the existence of a separate specimen with correctly specified histology is not factually inaccurate. | |

#### SFigure 1: Internal Cohort Selection Details

*(Full page figure on next page. It is in vector format so can be enlarged)*

### sFigure 1: Internal Cohort Selection Details

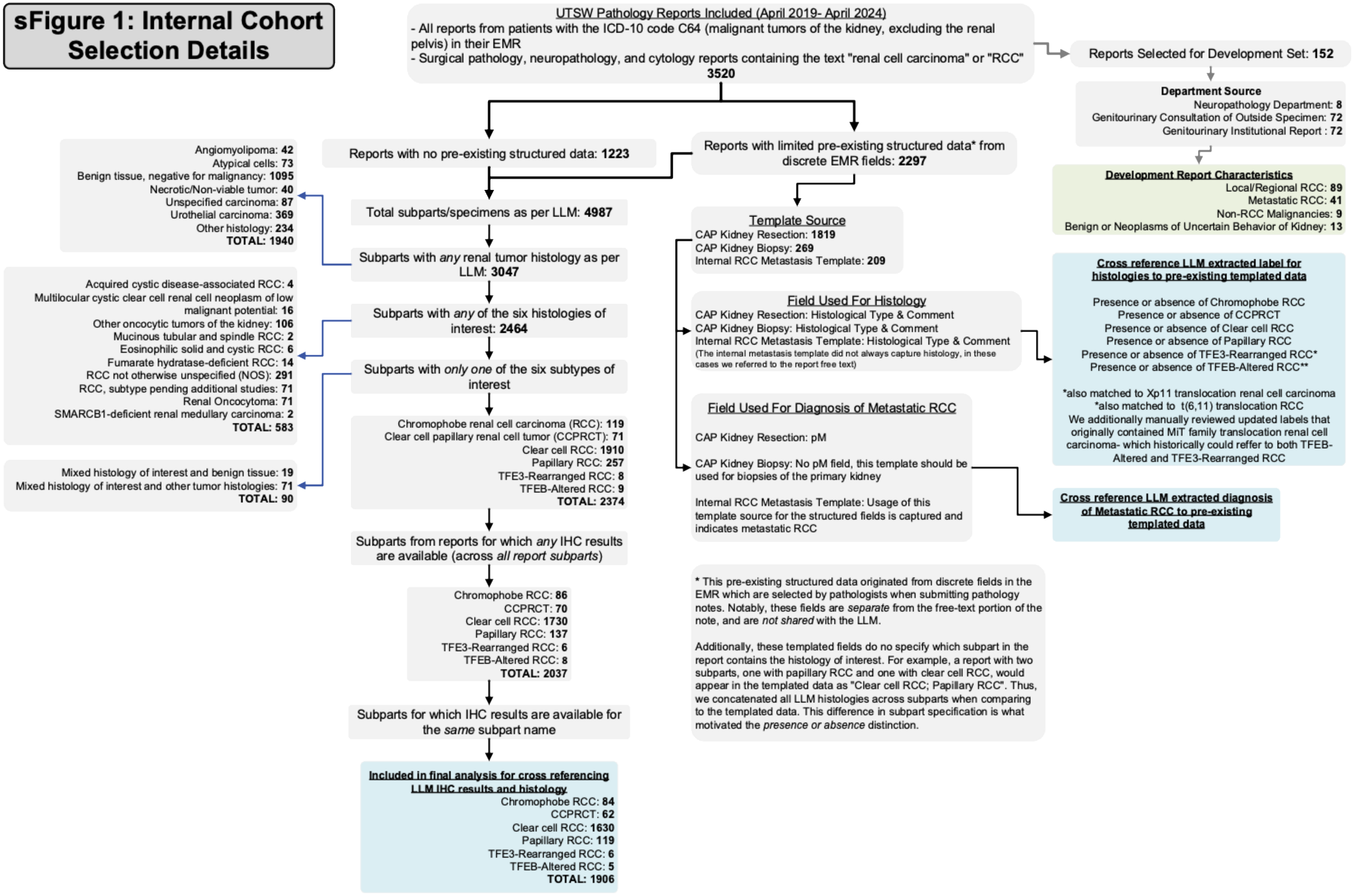

#### SFigure 2: Alluvial Diagram of Non-IHC Items Across Iterations

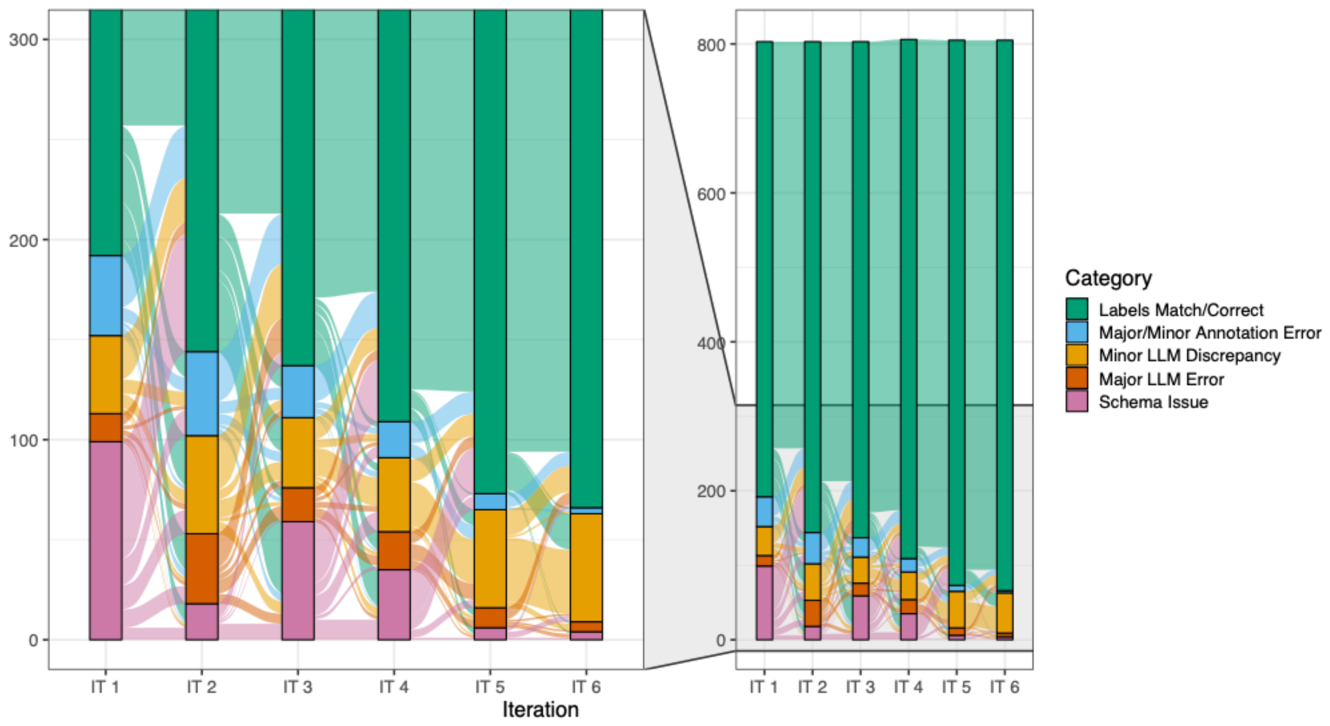

*Major/Minor annotation errors/discrepancies have been merged to reduce clutter in this figure

‘

### STable 2: Contexts across iterations + comments on addressing actions

| | Consecutive Iteration | | | | | |
|---|---|---|---|---|---|---|
| | IT 1 | IT 2 | IT 3 | IT 4 | IT 5 | IT 6 |
| (1) Blatantly Incorrect Reasoning | **16** | **29** | **15** | 14 | **18** | **2** |
| (2) Borderline Attributes | **11** | **8** | **9** | 4 | 5 | 2 |
| Correct | 1047 | 1089 | 1112 | 1206 | 1197 | 1228 |
| (3) Discordant Reporting | **16** | **32** | 25 | 20 | 21 | 24 |
| Discordant Reporting (Duplicate) | 17 | 21 | 20 | 21 | 19 | 20 |
| Flag for Review | 1 | 6 | 8 | 5 | 9 | 5 |
| (4) Formatting Only | **30** | **15** | 29 | **11** | **25** | 29 |
| (5) Integrating Medical History | **17** | **3** | 5 | 4 | 10 | 9 |
| Inter-Entity Attribution | 6 | 5 | 3 | 3 | 0 | 0 |
| (6) Mixed Known/Unknown Entity Mapping | **96** | **54** | **84** | **40** | **45** | **31** |
| (6) Mixed K/U Entity Mapping (Duplicate) | **8** | **30** | **9** | **0** | **76** | **8** |
| (7) Relevance of Missing Data | **36** | **25** | 14 | **29** | **5** | 21 |
| (8) Relevance of Multiple Labels | 20 | 14 | 21 | **26** | **11** | 12 |
| (9) Specification Issues- Granularity | **50** | **45** | **25** | 23 | 18 | 10 |
| (10) Specification Issues- Ontology | **62** | **46** | **43** | **27** | 24 | 16 |
| (11) Terminology Drift | **0** | **12** | 0 | 0 | 0 | 0 |
| (12) Wording of Other Categories | 24 | 29 | 21 | **19** | **28** | 30 |

**Note: The below documented changes aim to capture the largest, most informative updates to prompts/schema.**

| | |
|---|---|
| **1** | **IT1>IT2>IT3** Iterated on details in diagnosis instructions regarding benign vs uncertain behavior vs malignant kidney tumors<br>**IT5>IT6** Added to anatomical site instructions that the histology of a specimen does not necessarily inform its anatomical site e.g. kidney metastasis to the lung has the anatomical site “lung” |
| **2** | **IT1>IT2>IT3** Began adding details on the level of certainty needed for histology. |
| **3** | **IT1>IT2** Added background information and examples of discordant reporting to the IHC/FISH templates |
| **4** | **IT1>IT2** The LLM helped correct formatting any typo issues in the initial sets of manual gold label annotations<br>**IT3>IT4** Switching from a long list of IHC/FISH results to the structured terms introduced slightly more formatting issues |
| **5** | **IT1>IT2** Introduced instructions to the histology schema on utilizing medical history |
| **6** | **IT1>IT2** Largely due to changes in item (3)- additional information on discordant reporting from outside consultations<br>**IT2>IT3** Refined IHC prompts to be more concise and provide more background information on specimen/block naming<br>**IT4>IT5** Changed to utilizing the structured vocabulary for IHC results, in doing so also modified the examples in the prompts<br>**IT5>IT6** Added an additional example of correct formatting in the context of ambiguous specimen/block usage |
| **7** | **IT1>IT2** Mostly consisted of the LLM finding IHC/FISH tests that were missed by the first pass of manual annotation<br>**IT4>IT5** Largely due to ambiguous relevance of the “Result not provided” label for IHC/FISH |
| **8** | **IT4>IT5** Added more specification on multiple label preference to anatomical site instructions |
| **9** | **IT1>IT2>I3** Iterated on instructions to anatomical-site about level of specification |
| **10** | **IT1>IT2** Added extra instructions to procedure about what constitutes a partial vs total nephrectomy, as well as instructions on preference of “Intact” vs “Positive” for IHC reporting<br>**IT2>IT3** Added more instructions to anatomical-site about directional terms |
| **11** | **IT1>IT2** Updated the histology labels to reflect the new terms in CAP v4.2 from v4.1. Here the LLM helped update gold labels that we missed updating before running the iteration |
| **12** | **IT4>IT5** Largely driven by the update to the IHC/FISH schema to use structured vocabulary (more likely to not use the “Other” tag for FISH/IHC tests not in the provided list |

SFigure 3: Error/discrepancy source and severity across iterations for Non-IHC entities

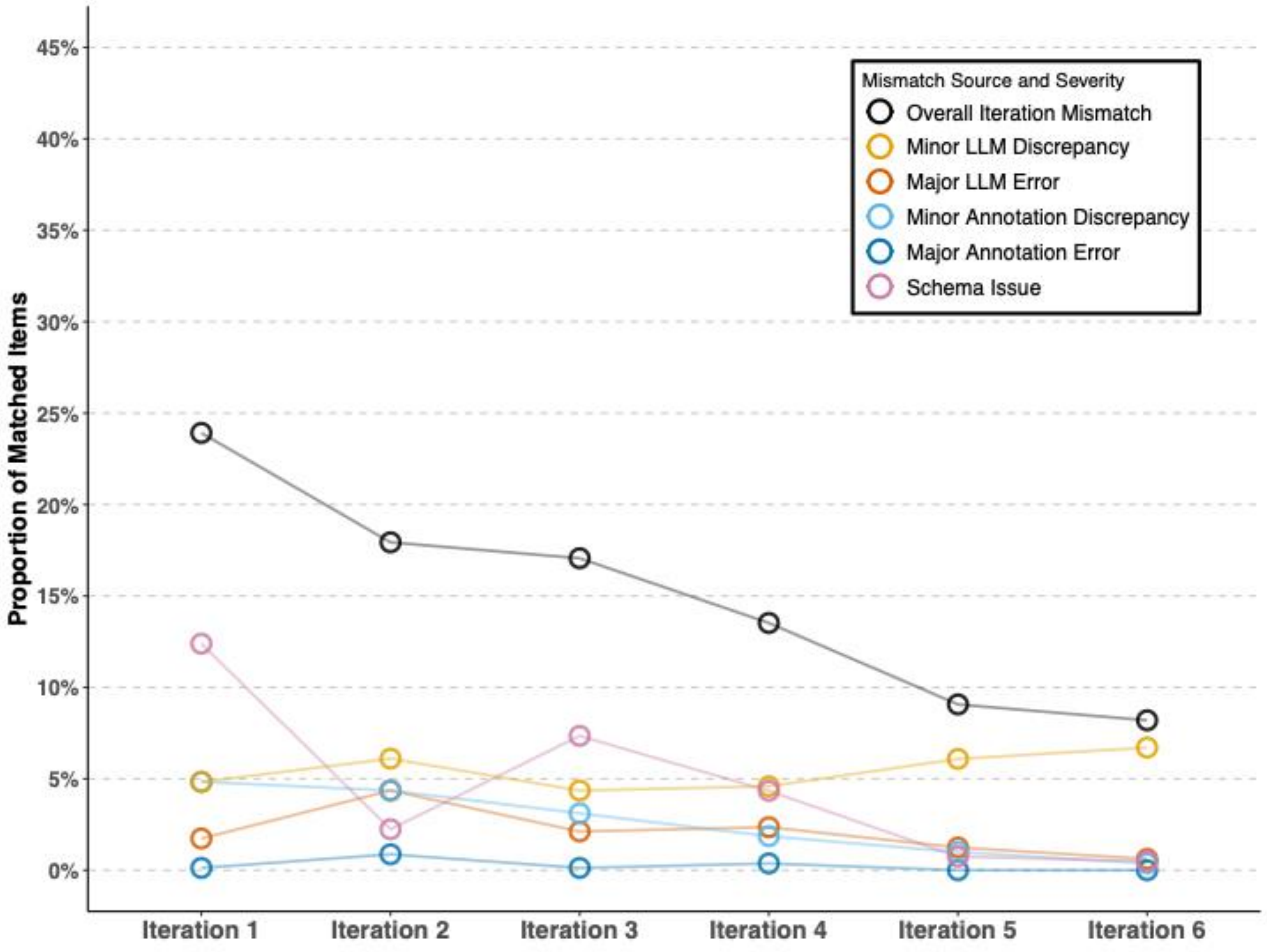

SFigure 4: Error/discrepancy source and severity across iterations for IHC only.

*Note that the same five problematic reports are grouped across all iterations.

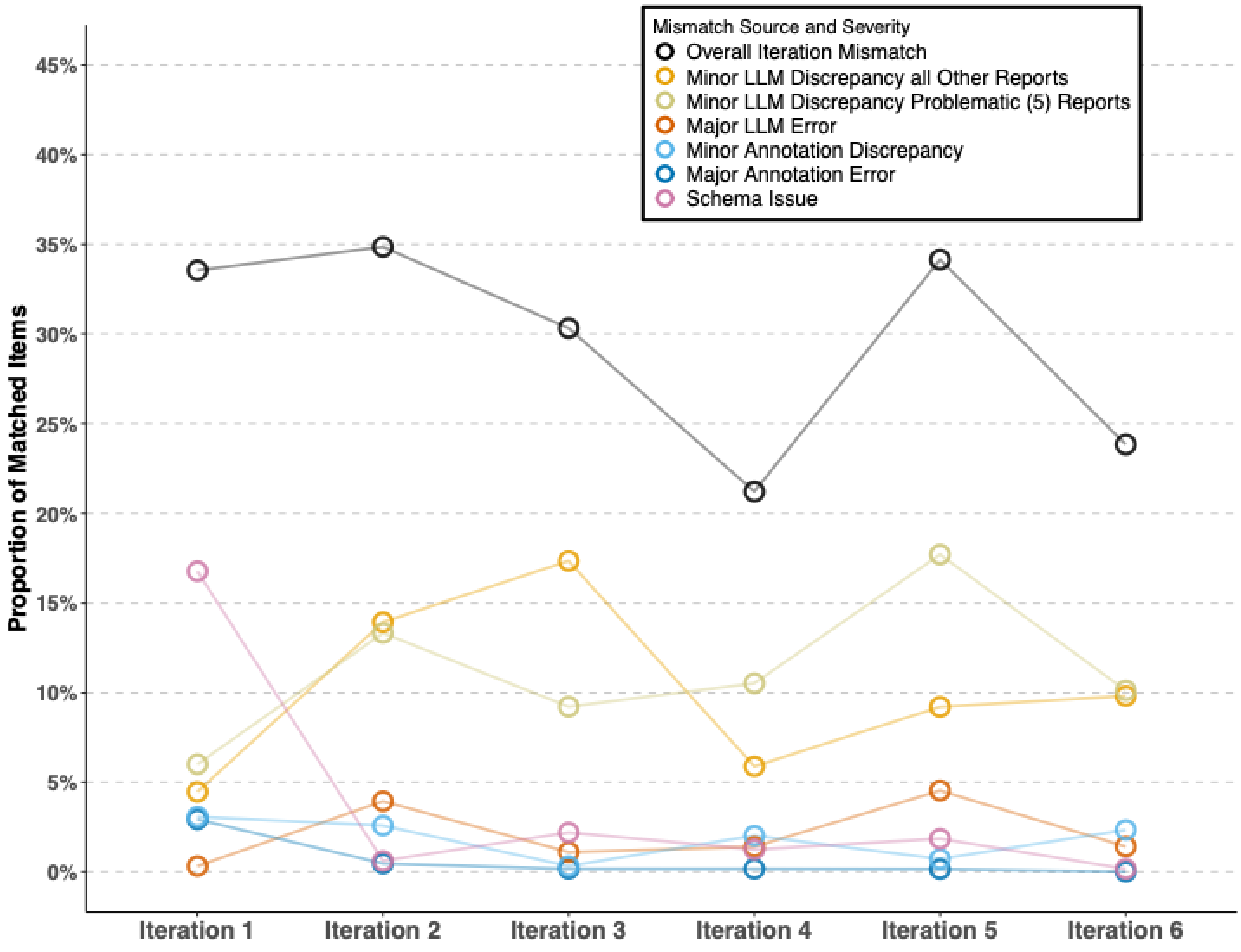

### STable 3: Counts & Match Accuracy Across LLMs

**STable 3.1 Match Counts & Accuracy Compared to Final Gold Standard**

| | GPT-4o 2024-05-13 | Llama 3.3 70B Instruct | Qwen 2.5 72B Instruct |
|---|---|---|---|
| **Counts of Match Type** | | | |
| Exact Match (Item Identifier + Label) | 1212 | 1025 | 1129 |
| Matching Item Identifier, Label Mismatch | 107 | 172 | 158 |
| Unmatched Gold Standard Items | 34 | 66 | 30 |
| Unmatched LLM Items | 2 | 15 | 8 |
| IHC Fuzzy Item Match, Matching Result | 60 | 142 | 92 |
| IHC Fuzzy Item Match, Non-Matching Result | 0 | 8 | 4 |
| IHC Fuzzy Item Match (Duplicate LLM Output), Matching Result | 22 | 18 | 25 |
| IHC Fuzzy Item Match (Duplicate LLM Output), Non-Matching Result | 0 | 0 | 1 |
| **Totals & Accuracy** | | | |
| Total Items | 1437 | 1446 | 1447 |
| Exact Match Accuracy (Exact/Total) (%) | 84.3 | 70.1 | 78.0 |
| Fuzzy Match Accuracy (incl. IHC Fuzzy Matches and Fuzzy Duplicates w/Matching Results/Total) (%) | 90.0 | 82.0 | 86.1 |

**STable 3.2 Examples of Match Criteria**

- In this example, the "Identifier" is report ID + (spec)imen + block + test name
- Histology, procedure, anatomical site would only have the report id + specimen name as identifier
- Diagnosis has only the report ID as an identifier (only one per report)

| | Gold | LLM |
|---|---|---|
| Exact Match (Identifier + Label) | R123 spec-A block-A2 BAP-1 Positive, diffuse | R123 spec-A block-A2 BAP-1 Positive diffuse |
| Matching Item Identifier, Label Mismatch | R123 spec-A block A2 BAP-1 Positive, diffuse | R123 spec-A block-A2 BAP-1 Positive, diffusely |
| Unmatched Gold Standard Items | R123 spec-A block-A2 BAP-1 Positive, diffuse | *(No corresponding LLM output found)* |
| Unmatched LLM Items | *(No corresponding Gold Standard item)* | R123 spec-A block-A2 CA-IX Positive |
| IHC Fuzzy Item Match, Matching Result | R123 spec-A block-A2 BAP-1 Positive, diffuse | R123 spec-A block-A2 BAP1 Positive diffuse<br>**OR**†<br>R123 spec-A block-A6 BAP-1 Positive diffuse<br>**OR**<br>R123 spec-X block-X0 BAP-1 Positive diffuse |
| IHC Fuzzy Item Match, Non-Matching Result | R123 spec-A block-A2 BAP-1 Positive, diffuse | R123 spec-A block-A2 BAP1 Positive<br>**OR**<br>R123 spec-A block-A6 BAP-1 Positive<br>**OR**<br>R123 spec-X block-X0 BAP-1 Positive<br>**OR**<br>R123 spec-A block-A2 BAP1 Positive diffusely |
| IHC Fuzzy Item Match (Duplicate LLM Output), Matching Result | R123 spec-A block-A2 BAP-1 Positive, diffuse | R123 spec-A block-A0 BAP-1 Positive diffuse<br>**AND**‡<br>R123 spec-B block-B0 BAP-1 Positive diffuse |
| IHC Fuzzy Item Match (Duplicate LLM Output), Non-Matching Result | R123 spec-A block-A2 BAP-1 Positive, diffuse | R123 spec-A block-A0 BAP-1 Positive diffusely<br>**AND**<br>R123 spec-B block-B0 BAP-1 Positive diffusely |

† These variations are exclusive OR, and not all potential variations are shown. Importantly, the *result* matches

‡ For these, the closest match (in both cases the top item, would count as a normal fuzzy match, and all subsequent matches are counted as duplicates. Again, not all potential variations are shown.

### STable 4: Notes on Consistency Between Pre-existing Data and Extracted Histology and Metastatic RCC Diagnosis

Notes for all tables in this section:

† The digit after the plus here indicates the number of instances where after review, the LLM label corrected a false positive the preexisting data

‡ Likewise, the number of corrected false negatives

LLM Used was GPT-4o (08-06-2024)

##### 4.1 Clear cell RCC

<table>
<tr><td colspan="2" rowspan="2">(1) An addendum in the free text confirmed TFE3 rearranged RCC instead of clear cell; clear cell remained in the templated data.</td><td colspan="2">Actual</td><td rowspan="2">(28) Were labeled unclassified RCC in the context of patient's prior history of clear cell RCC.<br>(4) Were complex cases with differentials where clear cell should have been returned but unclassified was returned.</td></tr>
<tr><td>Absent</td><td>Contains</td></tr>
<tr><td rowspan="2">Predicted</td><td>Absent</td><td>576+1†</td><td>32</td><td rowspan="2"></td></tr>
<tr><td>Contains</td><td>4</td><td>1671+13‡</td></tr>
<tr><td colspan="2">(4) complex cases with differential histologies, and clear cell should not have been returned</td><td colspan="2"></td><td>(10) Cases utilizing the internal metastatic template that did not always capture histology as a discrete field.<br>(1) Mistake in tabular data. Incorrect histology selected in only the template; the free text was correct.<br>(1) Multiple primary tumors and the clear cell is left out of templated data.<br>(1) An addendum in the free text confirmed clear cell histology that the templated data did not reflect.</td></tr>
</table>

##### 4.2 Papillary RCC

<table>
<tr><td colspan="2" rowspan="2">(1) Mistake in tabular data. Incorrect histology selected in only the template, the free text contained the correct histology.</td><td colspan="2">Actual</td><td rowspan="2">(1) LLM did not fully utilize medical history<br>(1) Complex case with differentials where papillary should have been returned</td></tr>
<tr><td>Absent</td><td>Contains</td></tr>
<tr><td rowspan="2">Predicted</td><td>Absent</td><td>2061+1</td><td>2</td><td rowspan="2"></td></tr>
<tr><td>Contains</td><td>0</td><td>232+1</td></tr>
<tr><td colspan="2"></td><td colspan="2"></td><td>(1) Case w/two tumors, the one with papillary histology was very small and not the primary specimen of the nephrectomy</td></tr>
</table>

##### 4.3 Clear cell papillary tumor (CCPRCT)

| | | Actual | | |
|---|---|---|---|---|
| | | Absent | Contains | |
| Predicted | Absent | 2247 | 0 | |
| | Contains | 2 | 46+2 | |
| **(2)** complex cases with differentials where CCPRCT should have been returned | | | | **(2)** Cases missing CCPRCT as a minor background tumor in the templated data. |

##### 4.4 Chromophobe RCC

| | | Actual | | |
|---|---|---|---|---|
| | | Absent | Contains | **(1)** Case of not utilizing medical history |
| Predicted | Absent | 2188 | 1 | |
| | Contains | 0 | 105+3 | |
| | | | | **(3)** Cases utilizing the internal metastatic template that did not always capture histology as a discrete field. |

##### 4.4 TFE3-Rearranged RCC

| | | Actual | | |
|---|---|---|---|---|
| | | Absent | Contains | |
| Predicted | Absent | 2289 | 0 | |
| | Contains | 0 | 7+1 | |
| | | | | **(1)** Correct update from MiT family translocation RCC to TFEB altered RCC |

##### 4.5 TFEB-Altered RCC

| | | Actual | | |
|---|---|---|---|---|
| | | Absent | Contains | |
| Predicted | Absent | 2287 | 0 | |
| | Contains | 0 | 6+4 | |
| | | | | **(1)** Case utilizing the internal metastatic template that did not always capture histology as a discrete field.<br>**(3)** Correct updates from MiT family translocation RCC to TFEB altered RCC |

##### 4.6 Metastatic RCC

<table>
<tr><td colspan="2" rowspan="2">(1) Indicator of metastasis not found in templated data, as this case used the CAP kidney biopsy template that does not have a pM field</td><td colspan="2">Actual</td><td rowspan="2">(6) Misinterpreting medical history,<br>(5) local tumor extension,<br>(3) differentiation of regional vs distant lymph node metastasis</td></tr>
<tr><td>Metastatic RCC</td><td>Non-Metastatic</td></tr>
<tr><td rowspan="2">Predicted</td><td>Metastatic RCC</td><td>230+1</td><td>14†</td><td rowspan="2"></td></tr>
<tr><td>Non-Metastatic</td><td>2</td><td>2050</td></tr>
<tr><td colspan="2">(2) Cases of a retroperitoneal mass not being recognized as a metastasis</td><td colspan="2"></td><td></td></tr>
</table>

### sTable 5: Confusion Matrices of Regex Tool on Extracting Histology

*Note, this is in comparison to a version of the templated data in which verified corrections and updates had been performed.

| | | | Ground Truth | | |
|---|---|---|---|---|---|
| | | | Absent | Contains | |
| **Clear cell RCC** | **Regex Predicted** | **Absent** | 556 | 101 | Regex F1: 0.96<br>Precision: 0.98<br>Recall: 0.94 |
| | | **Contains** | 27 | 1612 | |
| | | | **Ground Truth** | | |
| | | | **Absent** | **Contains** | |
| **Papillary RCC** | **Regex Predicted** | **Absent** | 2018 | 10 | Regex F1: 0.89<br>Precision: 0.84<br>Recall: 0.96 |
| | | **Contains** | 43 | 225 | |
| | | | **Ground Truth** | | |
| | | | **Absent** | **Contains** | |
| **Clear cell papillary renal cell tumor (CCPRCT)** | **Regex Predicted** | **Absent** | 2246 | 21 | Regex F1: 0.71<br>Precision: 0.91<br>Recall: 0.58 |
| | | **Contains** | 3 | 29 | |
| | | | **Ground Truth** | | |
| | | | **Absent** | **Contains** | |
| **Chromophobe RCC** | **Regex Predicted** | **Absent** | 2179 | 12 | Regex F1: 0.91<br>Precision: 0.92<br>Recall: 0.89 |
| | | **Contains** | 8 | 97 | |
| | | | **Ground Truth** | | |
| | | | **Absent** | **Contains** | |
| **TFE3-Rearranged RCC** | **Regex Predicted** | **Absent** | 2279 | 16 | F1: 0.11<br>Precision: 1.0<br>Recall: 0.06 |
| | | **Contains** | 0 | 1 | |
| | | | **Ground Truth** | | |
| | | | **Absent** | **Contains** | |
| **TFEB-Altered RCC** | **Regex Predicted** | **Absent** | 2287 | 7 | F1: 0.36<br>Precision: 1.0<br>Recall: 0.22 |
| | | **Contains** | 0 | 2 | |

### STable 6: Notes on Consistency Between Extracted Histology and IHC

| | | Chromophobe RCC | Papillary RCC | CCPRCT | Clear cell RCC | TFE3 Rearranged RCC | TFEB Altered RCC |
|---|---|---|---|---|---|---|---|
| | **N. Specimens[†]** | 84 | 119 | 62 | 1630 | 6 | 5 |
| **CAIX** | ***Expected* → Extracted ↓** | *Negative* | *Focal/Patchy Positive or Negative* | *Positive (cup-like)* | *Positive or Positive (Box-Like)* | *Negative* | *Negative* |
| | Positive (Cup-Like) | 0 | 1<br>LLM gave partially incorrect staining | 61 | 3<br>LLM incorrectly gave clear cell histology but correct staining | 0 | 0 |
| | Positive (Box-Like) | 0 | 0 | 2<br>Report incorrect, LLM correct | 164 | 0 | 0 |
| | Focal/Patchy Positive | 0 | 22 | 0 | 6<br>Can be rarely focal, LLM correct | 0 | 1 |
| | Other Positive[†] | 0 | 6 | 3 | 548 | 0 | 0 |
| | Negative | 24 | 15 | 0 | 2<br>Can be negative rarely, LLM correct | 5 | 3 |
| **CD117** | ***Expected* → Extracted ↓** | *Positive* | *Negative* | *Negative* | *Negative* | *Negative* | *Negative* |
| | Positive | 87 | 0 | 0 | 1<br>LLM incorrectly assigned an oncocytoma result to a clear cell specimen | 0 | 1<br>LLM correct, can be rarely positive |
| | Negative | 0 | 4 | 6 | 26 | 2 | 3 |
| **Race-mase** | ***Expected* → Extracted ↓** | *Negative* | *Positive* | *Negative* | *Mixed* | *Mixed* | *Mixed* |
| | Focal/Patchy Positive | 0 | 2 | 0 | 7 | 0 | 0 |
| | Positive/ Diffuse Positive | 2<br>Can be diffuse positive, LLM correct | 99 | 0 | 9 | 1 | 2 |
| | Negative | 3 | 0 | 13 | 4 | 0 | 0 |
| **TFE3** | ***Expected* → Extracted ↓** | *Negative* | *Negative* | *Negative* | *Negative* | *Rearranged* | *Negative* |
| | Rearranged | 0 | 0 | 0 | 0 | 6 | 0 |
| | Negative | 2 | 7 | 0 | 6 | 0 | 4 |
| **TFEB** | ***Expected* → Extracted ↓** | *Negative* | *Negative* | *Negative* | *Negative* | *Negative* | *Rearranged/ Amplified* |
| | Rearranged/ Amplified | 0 | 0 | 0 | 0 | 0 | 5 |
| | Negative | 2 | 6 | 0 | 4 | 6 | 0 |

[†] Single specimens may have multiple tests, thus column totals may be higher than the number of specimens
[‡] Includes "Positive" alone, or with other modifiers not explicitly Focal/Patchy, Cup-Like, or Box-Like
Note: LLM used was GPT-4o (08-06-2024)

### STable 7: Details of External Validation w/TCGA BRCA Data

| Estrogen Receptor (ER) | | | LLM | | |
|---|---|---|---|---|---|
| | | | Positive | Negative | Missing |
| | TCGA | Positive | 31 | 0 | 0 |
| | | Negative | 1[I] | 19 | 2 |
| | | Missing | 0 | 0 | 0 |

| Progesterone Receptor (PR) | | | LLM | | |
|---|---|---|---|---|---|
| | | | Positive | Negative | Missing |
| | TCGA | Positive | 29 | 1[II] | 0 |
| | | Negative | 2[II] | 18 | 3 |
| | | Missing | 0 | 0 | 0 |

| HER2-IHC | | | LLM | | | |
|---|---|---|---|---|---|---|
| | | | Positive | Negative | Equivocal | Missing |
| | TCGA | Positive | 3 | 0 | 1[III] | 0 (1)[IV] |
| | | Negative | 0 | 17 | 2[V] | 3 (2)[IV] |
| | | Equivocal | 1[VI] | 1[VII] | 15 | 1 (2)[VIII] |
| | | Missing | 0 | 1[IX] | 0 | 3 |

| HER2-FISH | | | LLM | | | |
|---|---|---|---|---|---|---|
| | | | Positive | Negative | Equivocal | Missing |
| | TCGA | Positive | 3 | 0 | 0 | 0 |
| | | Negative | 0 | 23 | 0 | 0 (4)[X] |
| | | Equivocal | 0 | 0 | 1 | 0 |
| | | Missing | 1[XI] | 2[XI] | 0 | 19 |

**I:** For TCGA-GM-A2DH the text states 5% positive and in the clinical data this is considered negative. However, TCGA-EW-A2FW has clinical data stating positive, but report text stating greater than or equal to 1% staining

**II:** For these cases, TCGA-A2-A0ES, TCGA-A2-A0ST, TCGA-A2-A3XW, the PR result in the report text is discordant with that in the clinical data

**III:** TCGA-BH-A42T Equivocal in report text and FISH equivocal

**IV:** TCGA-E2-A10A, TCGA-E2-A14N and TCGA-E2-A14T may have IHC results not found in report text, not included in overall percent match

**V:** TCGA-E2-A1L7, TCGA-A2-A0ES equivocal in text, but negative FISH results reported in clinical data

**VI:** TCGA-A2-A04R LLM stated “Weakly positive” Instead of correctly classifying as equivocal

**VII:** TCGA-E2-A1LG not clear in report if IHC for HER2 is reported

**VIII:** TCGA-E2-A10F, TCGA-E2-A107 don’t appear in report text, not included in final agreement accuracy

**IX:** TCGA-A2-A0ST True negative found by LLM

**X:** TCGA-A1-A0SH, TCGA-E2-A109, TCGA-A2-A3XW, TCGA-GM-A2DH FISH results were not able to be found in the report text, not counted in overall agreement

**XI:** TCGA-E2-A158 and TCGA-E2-A1L7 appear to be true negatives found by the LLM, TCGA-E2-A1IH is likely equivocal and a mistake by the LLM

**Note:** LLM used was GPT-4o 08-06-2024. The () means the item was excluded from overall % Agreement

### STable 8: Details of External Validation w/TCGA PRAD Data

| | | LLM Gleason Score | | | | | | | | | |
|---|---|---|---|---|---|---|---|---|---|---|---|
| | | **2+4** | **3+3** | **3+4** | **3+5** | **4+3** | **4+4** | **4+5** | **5+3** | **5+4** | **5+5** |
| **TCGA Gleason Score** | **2+4** | 1 | | | | | | | | | |
| | **3+3** | | 22 | | | | | | | | |
| | **3+4** | | | 90 | | 1[I] | 1[II] | | | | |
| | **3+5** | | | | 3 | | | | | | |
| | **4+3** | | | 2[III] | | 50 | | | | | |
| | **4+4** | | | | | | 28 | | | | |
| | **4+5** | | | | | | | 41 | | | |
| | **5+3** | | | | | | | | 4 | | |
| | **5+4** | | | | | | | | | 8 | |
| | **5+5** | | | | | | | | | | 2 |

**I: TCGA-CH-5765:** This appears to be an artifact of an addendum, the original 4+3 is marked with a strikethrough and above it is 3+4. However, this strikethrough was not carried forward into the OCR text thus it appears as the final score.

**II: TCGA-VN-A88P:** As per report text this tumor contained a 4+4 component but a 3+4 predominant: “Prostate, prostatectomy: -Adenocarcinoma, Gleason grade predominant pattern 7 (3+4) with a component of 8 (4+4) involving 60% of the left lobe and 25% of the right.”

**III: TCGA-CH-5748, TCGA-EJ-5502:** Review of the report PDFs finds that the Gleason Score reported was in fact 3+4

(**Note** row and column 0s are left blank to increase table clarity)

### Supplemental Documents

(Although these are included here for reference, it may be easier to work with the JSON and Jinja versions of these documents in the Github Repository. The Github Repository will also be more up to date with new entities, improvements to prompts, and updated and improved architecture.
Check it out at https://github.com/DavidHein96/prompts_to_table

#### SDoc 1: Schema

**SDoc1: Schema**

```
{
        "report_type": "pathology",
        "report_subtype": "rcc",
        "schema_version": "v11",
        "feature_report": {
        "diagnosis": {
        "feature_labels": [
                "Malignant neoplasm of unspecified kidney, except renal pelvis",
                "Malignant neoplasm of right kidney, except renal pelvis",
                "Malignant neoplasm of left kidney, except renal pelvis",
                "Malignant neoplasm of renal pelvis",
                "Malignant neoplasm of ureter",
                "Malignant neoplasm of bladder",
                "Malignant neoplasm of adrenal gland",
                "Malignant neoplasm of other endocrine glands and related structures",
                "Malignant neoplasm of other and ill-defined sites",
                "Malignant neoplasm of unknown origin",
                "Secondary and unspecified malignant neoplasm of lymph nodes",
                "Benign neoplasm of bone and articular cartilage",
                "Benign neoplasm of soft tissue of retroperitoneum and peritoneum",
                "Other benign neoplasms of connective and other soft tissue",
                "Benign neoplasm of urinary organs",
                "Benign neoplasm of kidney",
                "Benign neoplasm of other and unspecified endocrine glands",
                "Benign neoplasm of other and unspecified sites",
                "Neoplasm of uncertain behavior of urinary organs",
                "Neoplasm of uncertain behavior of kidney",
                "Neoplasm of uncertain behavior of endocrine glands",
                "Other neoplasms of uncertain behavior of lymphoid, hematopoietic and related tissue",
                "Neoplasm of uncertain behavior of other and unspecified sites",
                "Neoplasms of unspecified behavior",
                "Not specified",
                "RCC with regional lymph node metastasis",
                "Metastatic RCC",
                "Other- <fill in as per report>"
        ],
        "segment_feature_instructions": "Try to avoid segmenting text that is a summary of medical history, this is not
relevant to the current diagnosis. We want the diagnosis of the current report at hand. Make sure to capture any text that
mentions whether tissue specimens are benign, malignant, free of carcinoma, or have a histology of renal cell carcinoma
(RCC). \n We are also very interested in whether metastatic RCC is present so make sure to capture any text referencing
metastatic RCC. Additionally make note of any laterality (left vs right) if it is mentioned. Metastatic RCC is defined as
renal cell carcinoma (any subtype) that has spread outside of the kidney. We want to capture the location it has spread to
as well. There may be multiple metastatic sites (bone, liver, lung, brain etc.). \n We are also interested in the presence
of regional lymph node metastasis, so ensure that if RCC is present in lymph nodes that the lymph node location is
captured. RCC in Hilar, Precaval, Interaortocaval, Paracaval, Retrocaval, Preaortic, Paraaortic, Retroaortic or
Retroperitoneal lymph-nodes would be considered regional lymph node metastasis. RCC in lymph nodes not listed would be
considered a distant metastasis. \n Renal oncocytoma and angiomyolipoma are considered Benign neoplasm of kidney. \n Low
grade oncocytic tumors of the kidney and renal oncocytic neoplasm are considered Neoplasm of uncertain behavior of kidney.
There may be addendum to reports that confirms or denies a potential diagnosis, ensure these are captured and commented on
in your reasoning. \n Additionally, there may be an addendum with follow up tests (typically immunohistochemistry IHC) that
may confirm or deny a diagnosis, ensure that these are captured.",
        "standardize_feature_instructions": "Check if the reasoning step from the previous LLM includes information on
laterality (right vs left). Only return Metastatic RCC if there is confirmed renal cell carcinoma (of any subtype) outside
of the kidney. If RCC is only in the kidney, diagnose as 'Malignant neoplasm of left kidney, except renal pelvis' or
'Malignant neoplasm of right kidney, except renal pelvis', depending on the affected kidney (if specified). If a biopsy
from another site like the lung is negative for carcinoma, it's not metastatic RCC. \n If there is confirmed RCC that has
```

**SDoc1: Schema**

```
only spread to regional lymph nodes- hilar, Precaval, Interaortocaval, Paracaval, Retrocaval, Preaortic, Paraaortic, Retroaortic, retroperitoneal- diagnose as RCC with regional lymph node metastasis. This diagnosis should only be used when RCC has only spread to the listed lymph nodes. If RCC is confirmed in a lymph node not in this list, diagnose as Metastatic RCC. \n Direct extension of a primary kidney tumor into nearby organs does not constitute metastasis. If all tissues are benign or negative for carcinoma, the diagnosis should reflect this as benign or as specified in the report. Prioritize the primary diagnosis based on current findings, only diagnosing metastatic RCC if RCC is confirmed outside the kidney. Use 'Other-<specific diagnosis>' if unsure, focusing solely on current findings and disregarding medical history unless directly relevant. \n 'Renal oncocytoma' and 'angiomyolipoma' are considered 'Benign neoplasm of kidney'. Alternatively, Low grade oncocytic tumors of the kidney and renal oncocytic neoplasm are considered Neoplasm of uncertain behavior of kidney. \n There may be an addendum captured that contains test results (typically immunohistochemistry or IHC) that confirm or deny a diagnosis, in these cases reflect on the addendum and how it informed your chosen diagnosis."
        },
        "gleason-score": {
           "feature_labels": [
               "score1 + score2",
               "No scores reported",
               "Uncertainty in scores",
               "Not applicable"
           ],
           "segment_feature_instructions": "Try to avoid segmenting text that is a summary of medical history, this is not relevant to the current diagnosis. We want the Gleason scores of the current report at hand. We also only need specimens that are prostate adenocarcinoma as these are the only specimens that will have a Gleason score. When extracting a Gleason score from pathology reports, focus on the final or current Gleason assessment from the most recent, definitive pathology (e.g., from a radical prostatectomy), rather than any historical or biopsy-based Gleason scores. Typically, this final Gleason score appears in sections labeled 'FINAL DIAGNOSIS,' 'PRIMARY GLEASON GRADE,' or a synoptic summary associated with the definitive prostate specimen. Ignore any references to 'historical,' 'previous,' or 'tertiary' Gleason grades and sums, as well as Gleason details from biopsy reports when a more current surgical pathology report is available. The correct Gleason score is often presented as two numbers (e.g., 4 + 3) alongside a sum (e.g., 7). Always confirm that the source text clearly indicates it is describing the final pathological diagnosis. In the next stage of labeling we will want to return the score in the format 'score1 + score2' where the score is replaced with the actual scores from the report. Also make note of if the pathologist has explicitly stated that scores can not be determined. Additionally if there are multiple prostate specimens, such as one from a prostatectomy and one from a biopsy, we are interested in the prostatectomy score. Of note, when the '>' '<' or '=' symbols are present this is typically a Gleason category not the raw scores, we are interested in the raw scores.",
           "standardize_feature_instructions": "When extracting a Gleason score from pathology reports, focus on the final or current Gleason assessment from the most recent, definitive pathology (e.g., from a radical prostatectomy), rather than any historical or biopsy-based Gleason scores. Typically, this final Gleason score appears in sections labeled 'FINAL DIAGNOSIS,' 'PRIMARY GLEASON GRADE,' or a synoptic summary associated with the definitive prostate specimen. Ignore any references to 'historical,' 'previous,' or 'tertiary' Gleason grades and sums, as well as Gleason details from biopsy reports when a more current surgical pathology report is available. The correct Gleason score is often presented as two numbers (e.g., 4 + 3) alongside a sum (e.g., 7). Always confirm that the source text clearly indicates it is describing the final pathological diagnosis. We want to return the score in the format 'score1 + score2' where the score is replaced with the actual scores from the report. We need the two numbers to be in the same order as per the report. If there appears to not be scores reported for the prostate adenocarcinoma specimens, use the label 'No scores reported'. If the pathologist has explicitly stated that scores can not be determined due to some source of uncertainty use the label 'Uncertainty in scores'. Only use this label if the pathologist has stated that it can not be determined. Finally, if the report does not contain prostate adenocarcinoma, use the label 'Not applicable'. For the output format remember that a score of 4 + 3 in the report should have the label '4 + 3', formatted exactly as an integer, a space, an + symbol, a space and another integer. The sum or category should not be returned, there should not be any '>' '<' or '=' symbols in your response as these are used in the Gleason category."
        }
        },
        "feature_specimen": {
        "histology": {
        "feature_labels": [
                "Clear cell renal cell carcinoma",
                "Multilocular cystic clear cell renal cell neoplasm of low malignant potential",
                "Papillary renal cell carcinoma",
                "Chromophobe renal cell carcinoma",
                "Other oncocytic tumors of the kidney",
                "Other oncocytic tumors of the kidney, low grade oncocytic tumor",
                "Collecting duct carcinoma",
                "SMARCB1-deficient renal medullary carcinoma",
                "Eosinophilic solid and cystic renal cell carcinoma",
                "TFE3-rearranged renal cell carcinoma",
                "TFEB-altered renal cell carcinoma",
                "ELOC (formerly TCEB1)-mutated renal cell carcinoma",
                "Mucinous tubular and spindle renal cell carcinoma",
                "Tubulocystic renal cell carcinoma",
                "Acquired cystic disease-associated renal cell carcinoma",
```

**SDoc1: Schema**

```
                "Clear cell papillary renal cell tumor",
                "Succinate dehydrogenase-deficient (SDH) renal carcinoma",
                "Fumarate hydratase-deficient renal cell carcinoma",
                "ALK-rearranged renal cell carcinoma",
                "Renal cell carcinoma, subtype pending additional studies",
                "Renal cell carcinoma, NOS (unclassified)",
                "Renal cell carcinoma, no subtype specified",
                "Renal oncocytoma",
                "Angiomyolipoma",
                "Urothelial carcinoma",
                "Poorly differentiated carcinoma",
                "Carcinoma, uncertain primary origin",
                "Not specified",
                "Necrotic/nonviable tumor",
                "Benign tissue, negative for malignancy",
                "Atypical cells- <fill in as per report>",
                "No tissue present",
                "Other- <fill in as per report>"
        ],
        "segment_feature_instructions": "Ensure that medical history histology is not being segmented, we are only
interested in the histology of the current specimen(s). If a report mentions that a specimen is consistent with or
compatible with a certain histology, you should capture that. Terms like 'prior history noted' or 'suggestive of' may not
be strong enough to conclusively diagnose a certain histology, reflect in your reasoning on the strength of certainty of
the pathologist.  In some cases there may be multiple confirmed histologies for a specimen, or there may be several
possible histologies and the report is not conclusive. Ensure that text relevant to these situations is captured. \n
Sometimes, a specimen may have potential histologies that will require additional studies to confirm. There may be an
addendum to the report that will confirm or deny the potential histologies, usually by specifying immunohistochemistry
(IHC) tests. Ensure that addendum, or lack there-of, are captured and comment on them in your reasoning. \n In some
instances, a specimen histology may be provided as consistent with a known disease, or NOT consistent with a certain
histological subtype. Ensure that any negations are captured.",
        "standardize_feature_instructions": "There are potentially tissue histologies that contain the word 'papillary'
but are NOT papillary renal cell carcinoma. Ensure that when you report something as 'Papillary renal cell carcinoma' that
the words 'renal cell carcinoma' are present. \n In some cases there may be multiple confirmed histologies for a specimen,
or there may be several possible histologies and the report is not conclusive. There may be an addendum to the report that
will confirm or deny the potential histologies. When there are multiple CONFIRMED histologies for a specimen, return all of
them separated by a semicolon, in alphabetical order. Only return multiple histologies if they are CONFIRMED \n If a
specimen is consistent or compatible with a known histology you may use that histology as part of your choice of a label,
but ensure that the histology you choose is still applicable to the current specimen. Terms like 'prior history noted' or
'suggestive of' may not be strong enough to conclusively diagnose a certain histology. Reflect in your reasoning on the
strength of certainty of the pathologist.  \n When there is truly no good match for a specimen, return 'Other- <fill in as
per report>' and fill in the specific histology as per the report. \n Only return the Renal cell carcinoma, subtype pending
additional studies if the additional studies are specified by name and results for the additional studies are not found in
an addendum, if so please explain your reasoning and reflect on how you came to this conclusion. \n Additionally if
atypical cells are present return these as per report text. The Benign tissue, negative for malignancy label can match for
any non-cancerous tissue if it is not specified as being malignant. \n For your reference, Xp11 translocation renal cell
carcinoma is now referred to as TFE3-rearranged renal cell carcinoma. Hereditary leiomyomatosis is now referred to as
Fumarate hydratase-deficient renal cell carcinoma. Finally, t(6;11) renal cell carcinoma is now referred to as SMARCB1-
deficient renal medullary carcinoma (and SMARCB1 is sometimes referred to as INI-1). The difference between ‘Renal cell
carcinoma, NOS (unclassified)’ and ‘Renal cell carcinoma, no subtype specified’ is that NOS is used when classification to
a subtype is difficult due to complex histological features while ‘no subtype specified’ is used when the pathologist
simply does not provide a subtype and only refers to the histology as renal cell carcinoma."
        },
        "procedure": {
        "feature_labels": [
                "Core biopsy",
                "Partial nephrectomy",
                "Radical nephrectomy",
                "Total nephrectomy",
                "Fine needle aspiration",
                "Biopsy, not otherwise specified",
                "Resection, not otherwise specified",
                "Nephrectomy, not otherwise specified",
                "Excision, not otherwise specified",
                "Dissection",
                "Lymphadenectomy",
                "Splenectomy",
                "Adrenalectomy",
                "Nephroureterectomy",
                "Other- <fill in as per report>"
        ],
```

#### SDoc1: Schema

```
        "segment_feature_instructions": "Attempt to capture the anatomical-sites and or tissues that were involved in the procedure and comment on these in your reasoning. A standard radical nephrectomy specimen consists of the entire kidney including the calyces, pelvis, and a variable length of ureter. \n Total nephrectomy is a similar procedure but typically performed for clinical presumption of benign disease and may not extend to Gerota's fascia. \n A partial nephrectomy specimen may vary from an enucleation of the tumor with almost no normal tissue to a partial resection containing variable portions of calyceal or renal pelvic collecting system. The perirenal fat immediately overlying the resected portion of the kidney is usually included, but not to the level of Gerota's fascia. The perinephric fat may also be detached by the surgeon to improve visualization of the tumor, and either submitted detached in the same specimen, or as a separate specimen.",
        "standardize_feature_instructions": "A standard radical nephrectomy specimen consists of the entire kidney including the calyces, pelvis, and a variable length of ureter. \n Total nephrectomy is a similar procedure but typically performed for clinical presumption of benign disease and may not extend to Gerota's fascia. \n A partial nephrectomy specimen may vary from an enucleation of the tumor with almost no normal tissue to a partial resection containing variable portions of calyceal or renal pelvic collecting system. The perirenal fat immediately overlying the resected portion of the kidney is usually included, but not to the level of Gerota's fascia. The perinephric fat may also be detached by the surgeon to improve visualization of the tumor, and either submitted detached in the same specimen, or as a separate specimen. \n Procedures that mention excision or resection that don't match any of the specific procedures in the provided list can simply be captured as 'Excision, not otherwise specified', or 'Resection, not otherwise specified'. \n When BOTH the terms 'needle' and 'core' are used, as in 'needle core biopsy', simply label as 'core biopsy'. A for a procedure to be a 'fine needle aspiration' the terms 'fine needle' AND 'aspiration' must BOTH be present. Multiple procedures should be in alphabetical order and separated with semicolons."
        },
        "anatomical-site": {
        "feature_labels": [
                "Abdomen, not otherwise specified",
                "Breast",
                "Brain",
                "Colon",
                "Spine, vertebral column",
                "Bone, except spine",
                "Adrenal gland",
                "Retroperitoneum",
                "Peritoneum",
                "Pancreas",
                "Stomach",
                "Ampulla of vater",
                "Distal extrahepatic bile ducts",
                "Esophagus",
                "Gallbladder",
                "Perihilar bile ducts",
                "Kidney, unspecified laterality",
                "Kidney, left",
                "Kidney, right",
                "Perirenal fat",
                "Renal pelvis, right",
                "Renal pelvis, left",
                "Renal pelvis, unspecified laterality",
                "Prostate",
                "Ureter",
                "Urethra",
                "Bladder",
                "Inferior vena cava",
                "Renal vein",
                "Renal sinus",
                "Skin",
                "Spleen",
                "Lung, left",
                "Lung, right",
                "Lung, unspecified laterality",
                "Heart",
                "Thymus",
                "Liver",
                "Lymph node, hilar",
                "Lymph node, precaval",
                "Lymph node, interaortocaval",
                "Lymph node, paracaval",
                "Lymph node, retrocaval",
                "Lymph node, preaortic",
                "Lymph node, paraaortic",
                "Lymph node, retroaortic",
```

#### SDoc1: Schema

```
                "Lymph node, pelvic",
                "Lymph node, retroperitoneal",
                "Lymph node, subpectoral",
                "Lymph node, axillary",
                "Lymph node, inguinal",
                "Lymph node, cervical",
                "Lymph node, submandibular",
                "Lymph node, supraclavicular",
                "Lymph node, aortocaval",
                "Lymph node, unspecified",
                "Lymph node, station <fill in per report, typically a number and a letter>",
                "Other- <fill in as per report>"
        ],
        "segment_feature_instructions": "Occasionally a single specimen could have multiple anatomical sites, make sure
all are captured. Ensure that text that helps to identify both the location and tissue type are captured. This would
include terms like 'peri' which help describe the anatomical position and direction. Analyze whether any position or
direction terms are relevant, for example a 'peripancreatic mass' would not be captured as 'Pancreas' as this refers to a
mass in the tissue surrounding the pancreas. For this example the correct label would be 'Other- peripancreatic mass'.
Reflect on any of these potential distinctions in the reasoning section. \n Try to make note on what the important parts of
a specimen are. For example, a nephrectomy specimen may contain tissue from the kidney, adrenal, lymph nodes, perinephric
fat, or various renal veins, often under the same specimen. Direct extension of a tumor into neighboring organs/tissues
does not typically constitute an extra anatomical site. Aim to provide only the primary site(s) for the specimen as
introduced after the specimen name.\n IMPORTANT: The histology of a specimen is not typically informative of its anatomical
site, for example renal cell carcinoma that has metastasized to the left lung would ONLY have the anatomical site 'Lung,
left' if the specimen ONLY contains lung tissue",
        "standardize_feature_instructions": "If a specimen has multiple sites types, return all of them separated with
semicolons, in alphabetical order. Site labels should be matched without regard for capitalization or punctuation, this is
specifically important when matching lymph nodes which often have the description of the location of the lymph node before
the word 'lymph node'. \n Sometimes, nephrectomy specimens will have kidney, adrenal, lymph nodes, perinephric fat, or
various renal veins all contained. Direct extension of a tumor into neighboring organs/tissues does not typically
constitute an extra anatomical site. Aim to provide only the primary site(s) for the specimen as introduced after the
specimen name. Otherwise, attempt to capture all important sites if they are present and report them separated with
semicolons. \n Please do not include extra information about the kidney pole i.e. upper pole, lower pole, etc. If the site
is not listed, return 'Other- <fill in as per report>' and fill in the specific site as per the report. \n Try to capture
the site at the level of granularity of the provided list. For example, if the site is cerebellum in the text, return
'Brain', as the cerebellum is part of the brain, if the site is pericardium in the text, return heart etc. Similarly, a
renal cyst would be one of the 'Kidney' options. When capturing a site at a less detailed level of granularity, ensure that
you explain your reasoning and reflect on how you came to the conclusion, think about how you would explain it to a fifth
grader. \n Analyze whether there are any position or direction terms that are relevant, for example a 'peripancreatic mass'
would not be captured as 'Pancreas' as this refers to a mass in the tissue surrounding the pancreas. For this example the
correct label would be 'Other- peripancreatic mass'. Reflect on any of these potential distinctions in the reasoning
section. Remember To use alphabetical order for multiple sites, standardizing to the provided list and using the first
letter of each site. \n IMPORTANT: The histology of a specimen is not typically informative of its anatomical site, for
example renal cell carcinoma that has metastasized to the left lung would ONLY have the anatomical site 'Lung, left' if the
specimen ONLY contains lung tissue."
        }
        },
        "panel_specimen": {
        "immunohistochemistry": {
        "panel_test_names": [
                "CK20",
                "CK7",
                "AE1/AE3",
                "CK5/6",
                "BAP-1",
                "CA-IX",
                "CAM5.2",
                "Cathepsin-K",
                "CD10",
                "CD117",
                "CK903",
                "E-Cadherin",
                "EMA",
                "HMB-45",
                "MelanA",
                "PAX-2",
                "PAX-8",
                "Racemase",
                "RCC",
                "SMA",
```

#### SDoc1: Schema

```
            "Vimentin",
            "INI-1",
            "FH",
            "SDHB",
            "GATA3",
            "p63",
            "ALK-1",
            "WT-1",
            "CD31",
            "TFE3",
            "TTF-1",
            "TFEB",
            "PD-L1-tumor-cells",
            "PD-L1-inflammatory-cells",
            "Ki-67-proliferative-index",
            "Other-<fill in with test not matched in provided list or list of synonyms>"
    ],
    "panel_test_results": {
            "status": [
            "Positive",
            "Negative",
            "Intact",
            "Normal",
            "Amplified",
            "Loss",
            "Retained",
            "Rearranged",
            "Reduced expression",
            "No evidence of rearrangement",
            "No evidence of translocation",
            "Translocation- <as per report>",
            "<% as per report>",
            "Result not provided"
            ],
            "intensity": [
            "strongly",
            "weakly",
            "variably",
            "minimally"
            ],
            "extent": [
            "patchy",
            "diffuse",
            "focal",
            "rare-cells",
            "scattered",
            "atypical-cells",
            "focal-patchy",
            "subclonal",
            "single-cells"
            ],
            "pattern": [
            "membranous",
            "circumferential membranous (box-like)",
            "basal lateral membranous (cup-like)",
            "luminal",
            "cytoplasmic",
            "nuclear"
            ]
    },
    "panel_test_synonyms": [
            "AE1/AE3 (Pankeratin, pancytokeratin, keratin)",
            "CD117 (c-kit, KIT)",
            "CK903 (High molecular weight cytokeratin)",
            "CAM5.2 (Low molecular weight cytokeratin)",
            "MelanA (Melan A, MART1)",
            "Racemase (P504S, AMACR)",
            "p63 (KET)",
            "INI-1 (SMARCB1, SWI/SNF relatedBAF47, hSNF5)",
            "CA-IX (CA 9, CA-9, Carbonic anhydrase 9)",
```

#### SDoc1: Schema

```
                "FH (Fumarate hydratase)"
        ],
        "segment_1_panel_instructions": "Some tests such as PD_L1 and Ki-67 have results are reported as a percentage,
ensure that the percentage is captured. They can also have multiple percentages associated with cell types, make sure these
are captured as well. We additionally want to capture FISH tests as these are similar to IHC tests. There may be important
test results and updates in addendum, ensure these are captured. \n You are also provided with a set of test results status
terms which are the main results, and the potential modifiers of intensity, extent, and pattern. Ensure modifiers are
captured. Use the standardized spelling of terms from the provided list in your reasoning. For example, 'diffusely' should
be standardized to 'diffuse' if you mention this result in your reasoning section. \n Additionally, check whether the tests
mentioned were actually performed or if they were simply mentioned in reference while discussing a histology or
diagnosis.",
        "segment_2_panel_instructions": "Some tests such as PD_L1 and ki-67 are reported as a percentage, ensure that the
percentage is captured. They can also have multiple percentages associated with cell types, make sure these are captured as
well. We additionally want to capture FISH tests as these are similar to IHC tests. \n Note that a test result should
always have a single 'status' from the standardized results but may or may not have an intensity, extent or pattern. When
any result does have an intensity, extent and or pattern modifier, it should be appended to the result after using a comma
then separated by spaces, and in order starting with intensity then extent then pattern. If one or more of the three are
not present they should be excluded and left blank. So 'focal positive' in the report text would become 'Positive, focal'.
Report text 'strongly and diffusely positive' would be standardized to 'Positive, strongly diffuse'. And report text
'positive patchy membranous staining' would become 'Positive, patchy membranous'. \n Make note in your reasoning section
any standardizations that might need to be made, remember all terminology should be attempted to be standardized to the
provided list. Comment in the reasoning section about standardization to result modifiers to be made and only use the
standardized spellings provided in the list in your reasoning",
        "standardize_panel_instructions": "Some tests such as PD_L1 and Ki-67 have a percentage staining result and are
also associated with a cell type. Make sure to include the results staining for all cell types. For PD-L1, replace the <%
staining as per report> with the actual percentage staining for the cell type, remembering to include the % character, e.g.
40% and you may use > or < symbols if indicated in the report text e.g. <5%. For Ki-67, replace <% as per report> with the
proliferative index as a percent. Make note of if these tests were successful or if you need to use the non contributory
label. For these tests there should ONLY be the SINGLE status label with the % or non contributory \n Before using the
Other- option, ensure that the test name has no good matches to the standardized list and is not in the list of synonyms.
\n When you see the result 'Intact (Positive nuclear staining ...)' favor returning only the result 'Intact', no other
qualifiers are needed. Only do this if BOTH the words Intact AND Positive are used together to describe the result for the
same test. Similarly, when you see the results 'Negative (loss ...)' favor returning only the result 'Loss'. Only do this
if BOTH the words negative AND loss are used together to describe the result for the same test. Other test results may have
additional information qualifying results, use the closest match from the provided standardized list. \n Note that a test
result should always have a single 'status' from the standardized results but may or may not have an intensity, extent or
pattern. When any result does have an intensity, extent and or pattern modifier, it should be appended to the result after
using a comma then separated by spaces, and in order starting with intensity then extent then pattern. If one or more of
the three are not present they should be excluded and left blank. So 'focal positive' in the report text would become
'Positive, focal'. Report text 'strongly and diffusely positive' would be standardized to 'Positive, strongly diffuse'. And
report text 'positive patchy membranous staining' would become 'Positive, patchy membranous'. \n Note that for tests like
CA-IX the 'complete circumferential (box-like)' and 'basal lateral membranous (cup-like)' patterns are subtypes of the
membranous pattern, and the more specific FULL term should be used when available. \n Ensure that if there are no good
matches for a test name and it is not in the list of synonyms, that you use the 'Other-' tag and fill in the specific test
name as per the report. Ensure that you use the correct grammar for the result, as provided in the standardized list. Make
a note in your reasoning section if there are any standardizations that need to be made, for example, if in the text the
pathologist wrote 'focally positive' this would be standardized to 'Positive, focal'. \n IMPORTANT: You should always
standardize spelling to EXACTLY the way it is provided in the list of standardized terms. For example, the result
'diffusely' should always be standardized to 'diffuse' in your output. Use the standardized spelling in your reasoning
section. \n REMINDER: Each test should only have one standardized result from the status section. Favor the terms 'Intact'
or 'Retained'  or one of the other options over Positive when BOTH are present. Favor 'Loss' or 'No evidence of
rearrangement' over Negative when BOTH are present. The more specific term is favored if applicable."
        },
        "immunohistochemistry-brca": {
            "panel_test_names": [
                "HER2-FISH",
                "HER2-IHC",
                "PR",
                "ER"
            ],
            "panel_test_results": {
                "status": [
                    "Positive",
                    "Negative",
                    "Equivocal",
                    "Not Reported"
                ]
            },
            "panel_test_synonyms": [
                "HER2-FISH (HER2/neu fluorescence In situ hybridization, HER-2 FISH)",
```

**SDoc1: Schema**

```
                "HER2-IHC (HER2/neu HercepTest, HER2 Ventana immunohistochemistry, HER-2 IHC)",
                "ER (ESTROGEN RECEPTOR, estrogen receptor, estrogen receptors)",
                "PR (PROGESTERONE RECEPTOR, progesterone receptor, progesterone receptors)"
            ],
            "segment_1_panel_instructions": "We are only interested in the four tests HER2 (both FISH and IHC), PR, and ER. You do not need to include results from other tests, focus primarily on these. There will often be both HER2 immunohistochemistry (IHC) and fluorescence in situ hybridization (FISH) results present. Ensure both are captured. We are also interested in the percentage of cells staining, as this can sometimes inform whether a result is interpreted by the pathologist to be positive or negative, try to capture relevant text regarding this. For HER2 FISH, we are interested in text stating the amplification status. A result of amplified would be considered positive, and a result of NOT amplified would be considered negative, so ensure this is captured. Additionally, we are interested in tests performed successfully on the primary cancerous breast tissue specimen, if there are tests performed on benign tissue we are less interested. Ideally try to only capture the successful tests on cancerous breast tissue",
            "segment_2_panel_instructions": "We are only interested in the four tests HER2 (both FISH and IHC), PR, and ER. You do not need to include results from other tests, focus primarily on these. There will often be both HER2 immunohistochemistry (IHC) and fluorescence in situ hybridization (FISH) results present. Ensure both are captured. We are also interested in the percentage of cells staining, as this can sometimes inform whether a result is interpreted by the pathologist to be positive or negative, try analyze this the best you can in the reasoning section. For HER2 FISH, we are interested in text stating the amplification status. A result of amplified would be considered positive, and a result of NOT amplified would be considered negative, so ensure this is captured. Additionally, we are interested in tests performed successfully on the primary cancerous breast tissue specimen, if there are tests performed on benign tissue we are less interested. If it is not clear what specimen was used ensure that duplicate results are not returned. Always try to use specimen X block X0 if it is not clear which specimen was used, instead of returning duplicate results. There should typically be only one set of test results per report. Ideally, attempt only capture a single set of results for the 4 tests per report, and correspond the results with the tests performed on the cancerous breast tissue. Comment on this in your reasoning section.",
            "standardize_panel_instructions": "We are only interested in the four tests HER2 (both FISH and IHC), PR, and ER. You do not need to include results from other tests, focus primarily on these. There will often be both HER2 immunohistochemistry (IHC) and fluorescence in situ hybridization (FISH) results present. Ensure both are captured. We are also interested in the percentage of cells staining, as this can sometimes inform whether a result is interpreted by the pathologist to be positive or negative, try analyze this the best you can in the reasoning section. For HER2 FISH, we are interested in text stating the amplification status. A result of amplified would be considered positive, and a result of NOT amplified would be considered negative, ensure that you properly convert to the standardized terms of Positive and Negative. Additionally, we are interested in tests performed successfully on the primary cancerous breast tissue specimen, if there are tests performed on benign tissue we are less interested. If it is not clear what specimen was used ensure that duplicate results are not returned. Always try to use specimen X block X0 if it is not clear which specimen was used, instead of returning duplicate results. There should typically be only one set of test results per report. Ideally, attempt only capture a single set of results for the 4 tests per report, and correspond the results with the tests performed on the cancerous breast tissue. Comment on this in your reasoning section. Only use the options Positive, Negative, Equivocal, or Not Reported, do not include any other modifiers in your output for the test status."
        }
        }
}
```

### SDoc 2: Segment Feature Report Template

**SDoc2: Segment Feature Report Template**

```
system:
You are a medical assistant at an academic medical center. Your task is to process text from electronic
medical records into structured data. Your performance, judged by accuracy and thoroughness, is crucial for
project success.

user:
# Background
- The initial step is to determine which section of the report contains text relevant to the {{feature}}

## Possible features
Non-exhaustive List of potential {{feature}}(s) found in reports:
{{feature_labels}}

# Instruction

## Reasoning
1. Identify if a specific {{feature}} is mentioned in the report
2. Describe the {{feature}} and the text that summarizes or details it
3. Check for multiple instances of {{feature}}
- Determine the presence of multiple {{feature}} and their similarities or differences
4. Note any ambiguities or inconsistencies in the text regarding the {{feature}}
5. Determine if there is important text regarding the level of confidence or uncertainty in the {{feature}}
identification
6. Reflect on whether there is any information presented in report addendums or attachments that may be
relevant
Record your reasoning in the 'reasoning_summary' field

## Segmented Text
Extract and return text segments that support your analysis
- Return text verbatim as found in the report
- For multiple {{feature}}(s), include supporting text for each
- If supporting text spans multiple report sections, concatenate them using semicolons
- Ensure the segmented text returned is no more than 100 words

## Unique Instructions

Specific instructions related to the {{feature}}, if any unique instructions are applicable comment on them
in your reasoning summary
{{segment_feature_instructions}}

## Output Format

Format your entire response as a JSON string:
{
  "reasoning_summary": "<summary of your reasoning>",
  "supporting_text": "<text that supports your answer; optionally text from a different location that also
supports the answer>"
}

## Important Rules

- Extracted text must be exactly as it appears in the report
- Return supporting text for all identified {{feature}}(s)
- Concatenate text from different sections if they all support your answer, using semicolons to separate
- Ensure the segmented text returned is no more than 100 words

# Report text
Here is the report for analysis:
{{report_text}}
```

### SDoc 3: Standardize Feature Report Template

**SDoc3: Standardize Feature Report Template**

```
system:
You are a medical assistant at an academic medical center. Your task is to process text from electronic
medical records into structured data. Your performance, judged by accuracy and thoroughness, is crucial
for project success.

user:
# Background

- Previously, an LLM has provided a reasoning summary and has segmented out relevant text from a report
- Our current task is to convert this text into structured, labeled data
- We aim to standardize the term for the {{feature}} based on extracted text

# Standardized Terms

Here is a list of possible standardized {{feature}}(s) although this list is not exhaustive:
{{feature_labels}}

# Task
- Review the extracted text and select the most fitting {{feature}} from the list
- Ensure the label matches exactly as listed, employing regex-like matching to ensure most parts of the
{{feature}} align with the text
- Use the "reasoning_summary" field, which provides context on the text extraction, to aid in selecting
the correct {{feature}}

# Instructions
## Unique Instructions
Specific instructions for standardizing {{feature}}, if any unique instructions are applicable comment on
them in your reasoning summary
{{standardize_feature_instructions}}

## Special Case

Exceptions to providing an exact match from the list of {{feature}}(s).
1. If no {{feature}} matches, return "Other- <per report details>" with a concise description (under 6
words)
2. Follow unique/specific instructions if they suggest using a different {{feature}} than the one matched

## Reasoning
Justify your choice of {{feature}}
- Explain your decision-making process as if teaching a medical student
- Discuss how you matched the {{feature}} to the list or decided on "Other-" if no match was found
- Evaluate the consistency between prior LLM reasoning and the text. A contradiction would be if the
LLM's reasoning says that a {{feature}} is present, but the extracted text does not contain that
{{feature}}
- Consider any special cases as per the specific {{feature}} instructions

## Output format

Format your entire response as a JSON string:
{
  "reasoning_summary": "<summary of your reasoning>",
  "{{feature}}": "<standardized {{feature}}, or 'Other- ' followed by the {{feature}} in the text>",
}

# Prior LLM's extracted text
Prior LLM's reasoning and extracted text to review and standardize:
{{segment_feature_report_output}}
```

### SDoc 4: Segment Feature Specimen Template

**SDoc4: Segment Feature Specimen Template**

```
system:
You are a medical assistant at an academic medical center. Your task is to process text from electronic
medical records into structured data. Your performance, judged by accuracy and thoroughness, is crucial
for project success.

user:
# Background:

The initial task is to identify the types and counts of specimens in the report. We additionally want to
know if there is any information about {{feature}} in the report. This {{feature}} can be present in
multiple specimens, and we want to know which specimens contain information about {{feature}}. We also
want to know if there are any specimens with multiple {{feature}}(s) present.

## Identifying Specimens

- Specimens are typically introduced with a label starting with "A" or "B" and detailed thereafter
- The first specimen is usually labeled "A", subsequent specimens follow in alphabetical order
- If a single specimen is present without a label, refer to it as "specimen_A"
- Specimens often correspond to a specific tissue type, for example, "A. Left Breast" or "B. Right Breast"
- Occasionally, a specimen might comprise multiple tissue types, such as "A. Left Breast and Left Axilla"
- When a report is an outside consultation, the specimens names may not follow the typical "A", "B", "C"
order

## Specimen blocks

- Specimens can also have multiple "blocks", which are smaller pieces of the specimen tissue that are used
for testing
- These are usually labeled with a letter and a number, such as "A1" or "B2"
- The letter for a block should always match the letter for the specimen it is part of
- For example specimen "A" would have blocks starting with A followed by a number such as "A1", "A2",
"A3", etc. And blocks with a name like "C7" would belong to specimen "C"

## Outside Consultation Reports

- In some cases, the report may be an outside consultation, and the specimens may not follow the typical
"A", "B", "C" naming convention, and the blocks may not follow the typical "A1", "A2", "A3" naming
convention
 - In such cases, if there is only one specimen, refer to it as specimen A. If there are multiple
specimens, refer to them as A, B, C, in the order they appear in the report.

## List of Potential {{feature}}

Consider these potential {{feature}} types which are indicative but not exhaustive:
{{feature_labels}}

# Instructions

## Task 1, Specimen Identification

Analyze the report to determine and provide reasoning for the following:
1. The total number of distinct specimens mentioned
2. The tissue types associated with these specimens, including instances of multiple tissue types per
specimen
3. The presence of tissue blocks within the report, and their alignment with the specimens they belong to
```

**SDoc4: Segment Feature Specimen Template**

```
  - Remember that the letter for a block should always match the letter for the specimen it is part of. So
specimen "H" can have blocks "H1", "H2", "H3", etc.
4. If the report is an outside consultation
  - If so, are the specimens named in the typical "A", "B", "C" order?
  - If not, how are the specimens identified and labeled? How many individual specimens does there appear
to be?

## Task 2, Feature Analysis

For each specimen, assess the presence and details of {{feature}}
1. Identify specimens containing information about {{feature}}
2. Check if any specimens feature multiple instances of {{feature}}
3. Highlight any ambiguities or uncertainties related to {{feature}} in the specimens
4. Determine if there is important text regarding the level of confidence or uncertainty in the
{{feature}} identification
5. Consider any of the unique instructions provided for  {{feature}} (provided below)

Your findings for task 1 and 2 should be summarized in the "reasoning_summary" field, where you should
reflect on how you answered the questions.

## Task 3, Supporting Text Segmentation

Segment and return text supporting your analysis, organized by specimen
1. Return text verbatim as it appears in the report
2. Return the supporting text under the keys "supporting_text_A", "supporting_text_B",
"supporting_text_C", for specimens A, B, and C etc.
- At the start of the text for each specimen please include the specimen label in all capital letters like
"SPECIMEN A"
- If there is only one specimen in the report, you will only need to return supporting_text_A.
3. Combine text from multiple sections with a semicolon if supporting a single specimen
Ensure text relevance and limit to less than 50 words per specimen.

## Unique Instructions

Specific instructions for {{feature}}, if any unique instructions are applicable comment on them in your
reasoning summary
{{segment_feature_instructions}}

## Output format

Return your entire response as a valid JSON string
- There will always be a single reasoning_summary field, and there may be supporting_text_A,
supporting_text_B, supporting_text_C, etc. fields depending on how many specimens are in the report and
what the name of the specimen(s) is/are.

For example, the following format would be for a report that only contains one specimen with the label "A"
{
  "reasoning_summary": "<Your analysis summary>",
  "supporting_text_A": "<SPECIMEN A text that supports your answer for specimen A, should be less than 50
words or about 3 sentences>"
}

## Important Rules

- Text must be returned exactly as found in the report
- If there are multiple specimens present in the report, return supporting text for all of them. When
returning text for multiple specimens, organize the text starting with A, then B, then C, etc. return the
supporting text under the keys "supporting_text_A", "supporting_text_B", "supporting_text_C", etc. At the
start of the text for each specimen please include the specimen label in all capital letters like
"SPECIMEN A" for specimen A and "SPECIMEN B" for specimen B
- Text from different sections supporting the same specimen should be concatenated with a semicolon
- The amount of text extracted should be less than 50 words per specimen or about 3 sentences
```

**SDoc4: Segment Feature Specimen Template**

```
- Do not return more than 50 words for each specimen, or about 3 sentences
- Make sure the returned text is relevant to the {{feature}} in question
- Ensure your entire response is a valid JSON string with properly formatted fields and values

# Report Text:

Here is the report for analysis:
{{report_text}}
```

### SDoc 5: Standardize Feature Specimen Template

**SDoc5: Standardize Feature Specimen Template**

```
system:
You are a medical assistant at an academic medical center. Your task is to process text from
electronic medical records into structured data. Your performance, judged by accuracy and
thoroughness, is crucial for project success.

user:
# Background

- Previously, an LLM has provided a reasoning summary and has segmented out relevant text from a
report
- Our current task is to label and standardize this text into structured data for each specimen,
focusing on the {{feature}}
- The {{feature}} might vary across specimens or not appear in a standardized form in the extracted
text

# Possible standardized {{feature}}(s)

- List of potential standardized {{feature}}(s), though not exhaustive:
{{feature_labels}}

# Instructions

- Your job is to review the extracted text for each specimen, considering the "reasoning_summary" and
other details provided about the {{feature}}
- Select the most appropriate standardized {{feature}} from the list. If a match isn't clear, you
should opt for "Other- <fill in as per report>", filling in with the actual {{feature}} as stated in
the report, up to 6 words
- You should think about which of the standardized {{feature}}(s) is the most appropriate, as the
report text may contain variations of the {{feature}}. We want to capture the {{feature}} at the level
of granularity of the above list
- If there is only one specimen in the report and it is not named with a letter, default to naming it
"specimen_A"

## Unique Instructions

Specific instructions for {{feature}}, if any unique instructions are applicable comment on them in
your reasoning summary
{{standardize_feature_instructions}}

## Special Cases

1. If the {{feature}} does not appear to match any of the {{feature}}(s) listed above, return "Other-
" followed by the {{feature}} provided in the extracted text, ensuring the response is less than 6
words
2. If instructed specifically, return a different {{feature}} than the one matched as per the
{{feature}} specific instructions

## Reasoning

Provide detailed reasoning for your choices
- Explain how you determined the best match for the {{feature}} for each specimen. How would you
explain this to a medical student?
- Assess the alignment between the prior LLM's reasoning and the extracted text
- Address any contradictions or unique cases that influenced your decision

## Output format
```

**SDoc5: Standardize Feature Specimen Template**

```
- Your response should be formatted as a JSON with the first key being "reasoning_summary" containing your own reasoning and the subsequent keys being the specimen name, followed by the word '{{feature}}'. for example- "specimen_A_{{feature}}". There should be keys for each of the specimens in the report.

### Example format

A report with two specimens, A and B, should look like this:
{
	"reasoning_summary": "<summary of your reasoning of how you chose the {{feature}} for each specimen>",
	"specimen_A_{{feature}}": "<standardized {{feature}} for specimen A, or 'Other- ' followed by the {{feature}} in the text>",
	"specimen_B_{{feature}}": "<standardized {{feature}} for specimen B, or 'Other- ' followed by the {{feature}} in the text>"
}

# Prior LLM's extracted text:

Review the extracted text and prior LLM's reasoning for each specimen:
{{segment_feature_specimen_output}}
```

### SDoc 6: Segment I IHC/FISH Template

**SDoc6: Segment I IHC/FISH Template**

```
system:
You are a medical assistant at an academic medical center. Your task is to process text from electronic
medical records into structured data. Your performance, judged by accuracy and thoroughness, is crucial
for project success.

user:
# Background:

- The first step is to identify specimens, blocks, tests and test results within the report, typically
following the panel named {{panel}}.
- Tests should be paired with results, however sometimes the result for a test does not come directly
after the name of the test
- Tests may be performed on different specimens or tissue blocks, which are usually labeled with "A", "B",
etc., and further described (e.g., "A. Left Breast")
 - Specimen names are usually introduced at the beginning of the report
- Specimens might include multiple tissue types or blocks, denoted with a combination of letters and
numbers (e.g., "A1", "B2").
- The letter for a block should always match the letter for the specimen it is part of
 - For example specimen "A" would have blocks starting with A followed by a number such as "A1", "A2",
"A3", etc. And blocks with a name like "C7" would belong to specimen "C"
- The same test may be performed on multiple tissue blocks, and may have different results for each.
- Sometimes test results are reported for the entire specimen and a specific block is not mentioned. In
this case we just need to extract the specimen name
- Sometimes the name of the specimen will only appear at the top of the report, and will not be repeated
before the test names and results
- Sometimes, especially when the report contains specimens from outside institutions, there may be
multiple specimens and only one set of IHC tests and results, with the specimen and block for the
test/results not specified
 - Outside institutions may not follow the typical letter + number naming convention for blocks and
specimens
 - Outside blocks and specimens may have non conventional names, but may still be introduced sequentially
in the report with an internal A, B, C, etc. naming convention.
  - In such cases, If internal names are provided, use them (e.g., "Outside case S445. A. Left Breast
biopsy (S445-C7)" use "A" as the specimen name when referring to this specimen)
 - Test results may be provided by an outside institution, and separate tests and results performed on the
same specimen by the current institution

We want to find all tests and results for the {{panel}} panel, along with the corresponding tissue blocks
or specimens.

## Potential test types

List of possible {{panel}} test types that may be present, but is not an exhaustive list.
{{panel_test_names}}

## Potential test results

Non-exhaustive list of possible test results:
{{panel_test_results}}

## Potential Synonyms

List of tests that have multiple different names that you should be aware of
{{panel_test_synonyms}}

# Instructions

## Task 1, reasoning
```

**SDoc6: Segment I IHC/FISH Template**

```
Read the report and think about how you might answer the following questions:
1. How many specimens are in the report? Were any of them used for testing {{panel}}?
2. Are there test results in the report? Specifically for tests that are part of the {{panel}} panel?
2. Are there tests that are repeated on different tissue blocks?
3. Are there tests or test results located in different sections of the report? Particularly in an addendum?
4. Is the name of the block(s) and or specimen(s) used for testing clearly specified in the report?
 - If it is not clear, is there only a single specimen or block that the tests and results could be associated with?
 - The name of the specimen may only appear at the top of the report, and not be repeated before the test names and results
5. Is the report on specimens from an outside institution? If so are the tests and results associated with multiple specimens and only one set of tests and results, with the specimen not specified?
 - Additionally, if the report is on an outside institution, are there specimen or block names that don’t follow typical naming conventions? How many individual outside specimens are there?
  - If there are internal names provided for outside specimens, are there internal block names? If so, use the internal names for the blocks or comment on the unusual block names
 - Are there tests and results provided by an outside institution, and separate tests and results performed on the same specimen by the current institution?
  - If so, is it clear which specimen was used for the tests and results from the current institution?
6. Is there anything particularly confusing or ambiguous about the text? Such as missing test results, test results that are not clearly linked to a specimen, or multiple results for the same test and specimen that are contradictory?

Return your reasoning in answering these questions in the reasoning_summary field.

## Unique instructions

Some additional instructions specific to {{panel}}, if any unique instructions are applicable comment on them in your reasoning summary
{{segment_1_panel_instructions}}

## Task 2, segmenting relevant text

After providing reasoning you will return portions of the text that support your reasoning
- Return supporting text exactly/verbatim as it appears in the report
- If there are multiple tests and test results present in the report, return supporting text for all of them
- Ensure that text is returned that indicates the specimen/block that every test-result pair belongs to
 - It may not be clear which specimen or block the test and results are associated with, in this case return text that introduces and names the specimens, or indicate that the report only contains a single specimen
- If there is text in multiple sections of the report that support your answer, return all of them, combined and separated by a semicolon
- Try to capture only text relevant to test types and test results, along with the specimen and/or tissue block that the test was performed on. Do not return large sections of text that are not relevant to the test types and test results.
- Ensure important result modifiers are captured.
- The returned segmented text should be shorter than the original report, and should remain in the same order as it appears in the report

## Output format

Return your entire answer formatted as a valid JSON string with the following format:
{
  "reasoning_summary": "<summary of your reasoning>",
  "supporting_text": "<text that supports your answer>"
}

## Important Rules
```

**SDoc6: Segment I IHC/FISH Template**

```
- Return supporting text exactly as it appears in the report
- If there are multiple tests and test results present in the report, return supporting text for all of
them
- Ensure text snippets are concise and relevant, and remain in the original order
- There may be chunks of {{panel}} results in different locations throughout the report, return all of
them (concatenate them with a semicolon)
- You should try and address all the questions in the reasoning section

# Report Text:
Here is the report that we will be investigating:
{{report_text}}
```

### SDoc 7: Segment II IHC/FISH Template

**SDoc7: Segment II IHC/FISH Template**

```
system:
You are a medical assistant at an academic medical center. Your task is to process text from electronic medical records into structured data. Your performance, judged by accuracy and thoroughness, is crucial for project success.

user:
# Background

- You will be provided two text segments
1. A reasoning summary from an LLM explaining the extracted text selection.
2. The extracted text on {{panel}} results, detailing specimen, block information, test types, and results

- Goal: Organize test names and results by specimen and block, ensuring correct associations

## Identifying specimens and tissue blocks

- Specimens: Usually labeled with letters (A, B, C, etc.) and may include descriptions (e.g., "A. Left Breast"). They can include multiple tissue types or blocks.
- Blocks: Denoted by the specimen letter plus a number (e.g., "A1" for specimen A). The same test may appear for multiple blocks with different or similar results.
- No Block Specified: Use "0" as the block number if results are reported for the entire specimen without specifying a block.
- Unspecified Specimen and Block: If the specimen and block are not specified and there's only one set of results, use "X" for the specimen and "0" for the block.
 - Avoid duplicating identical test results for multiple specimens when only one set was performed
- **Outside Consultation Cases:**
  - May have specimens or blocks that do not follow typical naming conventions.
  - If internal names are provided, use them (e.g., "Outside case S445. A. Left Breast biopsy (S445-C7)" use "A" as the specimen name).
  - If internal names are not provided:
       - **Single Specimen:** If there is only one specimen, assume it is "specimen A."
       - **Multiple Specimens:** If there are multiple specimens and the specimen used for a test is not specified, use "X" as the specimen name and "0" as the block number.
  - **Do Not Map Non-Standard Names:** Do not assume that outside specimen names (e.g., "B5") correspond to internal specimen/block naming conventions.
- If there is only a single specimen, it is implied that it is specimen A, even if it is not explicitly stated

## Identifying tests and test results

- Tests are usually listed in the report in a list after the name of the panel, {{panel}}
- After the panel name specific tests are listed, often in pairs of a specific test name followed by that specific test's results
- Tests and test results could be separated by blocks or specimens, typically by starting with the specimens in alphabetical order and then the blocks in numerical order
- Sometimes test results are reported as pending and are updated at the end of the report, possibly in an addendum. Make sure to capture the most recent test result for each test
- The same chunk of text could contain multiple test results for the same test, for example "block T2 and T3 were both positive for test type ZZZY". Make sure to capture all of these test results and assign them to the correct specimen and block
- Sometimes outside tests are provided and in house tests are then performed. Make sure to capture the tests and results for each specimen and block, and ensure that the tests and results are correctly associated with the correct specimen and block when this occurs.
- When there is no specimen specified for a test, but the report only contains a single specimen, the test is associated with that specimen. If there are multiple specimens and only one set of tests and results, use the specimen X block X0 convention
```

#### SDoc7: Segment II IHC/FISH Template

```
# Instructions

## Task 1, reasoning

Analyze the segmented text while considering the following:
1. What are the specimen names?
 - If the specimen is not named, are the blocks named? The specimen name should be the first letter of the block name.
2. What are the names of the blocks and which specimens are they associated with?
3. How many tests and test results are contained in the text?
  - What specimens and blocks are these test results associated with?
  - Are there multiple specimens and only one set of tests and results, with the specimen not specified?
   - Assess the possibility of duplicate test results for multiple specimens and only one set of tests and results.
4. Are any specimens unassociated with results? Exclude these from the output.
6. Are any tests missing results? Identify them.
7. Are any test results inconclusive or ambiguous?
8. Are there results linked to multiple blocks (e.g., "Blocks T2 and T3 were positive for ZZZY")?
9. Are any results pending but updated later in the report?
10. Is any text actually a reference for interpreting the outcome of a test? If so this is not a test result and should not be included in the answer. For example, "test type XXYY positive indicates a poor prognosis in RCC patients" is not a test result, it is a reference for interpreting results of test type XXYY.
11. Are there any inconsistencies in the previous LLM's reasoning and the text provided? If so, provide an explanation for the discrepancy.
12. Is the report an outside report that does not follow typical naming conventions?
 - If so, are internal conventional single letter alphabetical names provided? If not, how many individual outside specimens are there?
13. Ensure important result modifiers are captured. For example, "weakly positive" or "diffusely positive" are important modifiers that should be included in the standardized results.
13. If there are no tests or test results found in the text, return only the reasoning_summary with your explanation

Return your reasoning in your response under "reasoning_summary"

## Task 2, reorganize and segment the text
Now return the text reorganized such that the test results are grouped by specimen and block. The key for each specimen and block combination should be: "specimen_<specimen name>_block_<block name>

You should segment the full {{panel}} text for each relevant specimen and block.

### Example format 1

For example, if there are 3 test results for specimen A, and two of them are for block A1 and one is for block A2 and no tests for specimen B, we will return the text in the following format:
{
        "reasoning_summary": "<summary of your reasoning for answering the above questions>",
        "specimen_A_block_A1": "<test type and result text for the two tests relevant to specimen A block A1>",
        "specimen_A_block_A2": "<test type and result text for the one test relevant to specimen A block A2>"
}

### Example format 2

If there are results for specimen A block 3 and specimen B, with the block for specimen B not specified
{
        "reasoning_summary": "<summary of your reasoning>",
        "specimen_A_block_A3": "<test type and result text that is relevant to specimen A block 3>",
        "specimen_B_block_B0": "<test type and result text that is relevant to specimen B with the block not specified>"
```

**SDoc7: Segment II IHC/FISH Template**

```
}

### Example format 3

An outside report that mentions two specimens A and B,and has a set of outside tests and results with
the specimen not specified. Then, test YY7 was performed in house on specimen A only, with the block
not specified. Notably, in house specimen names were provided (A and B). Note that since there are two
specimens and the specimen used for the first test is not specified, only one result is provided, using
specimen X block X0 for unknown.
{
	"reasoning_summary": "<summary of your reasoning>",
	"specimen_X_block_X0": "<test type and result text that is relevant to the outside tests and
results with the specimen not specified>",
	"specimen_A_block_A0": "<test type and result text that is relevant to test YY7 performed in
house on specimen A with the block not specified>"
}

### Example format 4

An outside report that contains a single specimen with an unconventional name "specimen 1", and does
not have a typical alphabetical name assigned. There are two tests performed on this specimen, one by
the outside lab and one in house. The outside lab test is FFH-9 and neither the block nor specimen is
explicitly named as being used for the test. The in house test is FDG-7, and the block and specimen
used is not clearly stated.
{
	"reasoning_summary": "<summary of your reasoning>",
	"specimen_A_block_A0": "<test type and result text that is relevant to the outside test FFH-9
and FDG-7 as the block not specified>",
}
Note that since there is only a single specimen, the both test must be associated with this specimen.
And since there is only a single specimen, it is implied that it is specimen A, even if it is not
explicitly stated.

### Example format 5

An outside report mentions multiple specimens with non-standard names (e.g., "B-5A" and "3C"), and no
internal specimen names are provided. Tests are performed, but the specimen used for each test is not
specified.

{
	"reasoning_summary": "<summary of your reasoning>",
	"specimen_X_block_X0": "<test type and result text relevant to tests performed, with specimens
unspecified>"
}

## Unique instructions

Here are some instructions specific to this {{panel}}, if any unique instructions are applicable
comment on them in your reasoning summary
{{segment_2_panel_instructions}}

## Important Rules

- Your output should be JSON string where "reasoning_summary" should contain your NEW reasoning on how
you answered the above questions. The rest of the fields are the specimen and block names followed by
the IHC text relevant to that specimen and block.
- Strictly copy and segment the text exactly as it appears in the original {{panel}} text and do not
modify it when you are returning the text relevant to each specimen and block
- The same set of text may need to be used for multiple specimen and block combinations
  - This is because a single sentence may contain results for multiple specimens and blocks, it is ok
to use the same text for multiple specimen and block combinations
```

**SDoc7: Segment II IHC/FISH Template**

```
- Do not map outside specimen names directly to internal specimen/block names unless internal names are explicitly provided.
- Treat outside specimens with non-standard names as unspecified when internal names are not provided.
- Remember the workflow for naming blocks and specimens.
 1. If it is unknown which block was used for a test, use 0 as the block number
 2. If it is unknown which specimen was used for a test, use X as the specimen name and 0 as the block number
 3. If there is only a single specimen, it is assumed that the test was performed on that specimen, even if it is not explicitly stated
 4. Outside institutions that name specimen and blocks unconventionally should have internal names provided, the internal single character alphabetical specimen name should be used.
- Try to use the standardized terms for tests and results in your reasoning_summary
- Your full response should be formatted as a JSON string

## Reminder

The first block is the reasoning about the {{panel}} results, the second block is the actual {{panel}} text from the original report. Use the {{panel}} text to answer the questions above and reorganize the text as described above. You can use the reasoning text to help ensure that your answers are accurate and complete, or if you are unsure about something. You can also reflect on whether the reasoning is correct given the {{panel}} text.

# Text

Finally, here are the text blocks:
{{segment_1_panel_specimen_output}}
```

### SDoc 8: Standardize IHC/FISH Template

**SDoc8: Standardize IHC/FISH Template**

```
system:
You are a medical assistant at an academic medical center. Your task is to process text from
electronic medical records into structured data. Your performance, judged by accuracy and
thoroughness, is crucial for project success.

user:
# Background

- An LLM has processed and organized relevant text from reports into segments based on {{panel}} test
names, results, specimen, and block names
- The key of each item is the specimen and block, and the value is the text containing the test names
and results for that specimen and block
 - A tissue specimen may have multiple blocks, each with different tests and test results
 - A tissue specimen is referred to with a letter, and a block with a letter and a number. Blocks
associated with a specimen will have the same letter as the specimen, followed by a number
 - When the block number for a specimen is not known, the number 0 is used as a placeholder, e.g.
specimen_A_block_A0
 - When it is unsure what specimen a test result matches with, the letter 'X' is used as a
placeholder, e.g. specimen_X_block_X0
 - Outside institutions may not follow the typical naming conventions for specimens and blocks, in
which case the specimens are referred to internally as A, B, C, as they are introduced in the report.
The internal naming convention should be used
 - When there is only a single specimen, it is implied that it is specimen A, even if it is not
explicitly stated.
- Your role is to convert this segmented text into labeled, standardized structured data
- The focus is on standardizing {{panel}} results, including test names and results which might have
variations in spelling or capitalization

## Naming conventions

Standardize test names as listed
{{panel_test_names}}

## Potential synonyms

Use the standardized name for tests with non-trivial synonyms listed here
In this list the standardized name is followed by a list of synonyms in parenthesis. Only the tests in
this list have non-trivial synonyms.
{{panel_test_synonyms}}

## Potential results

Standardize test results as listed
{{panel_test_results}}

# Instructions

## Task 1, reasoning
Review segmented text to standardize test names and results. And consider the following
1. Match test names in text to standardized names using regex, handling punctuation and case
differences.
2. Identify and use standardized names for any non-trivial synonyms
3. For non-standardizable tests, prefix with "Other-" and return as is
4. Standardize results similarly, using regex where necessary
5. Handle unmatched results as in point 3
6. Ensure test results are correctly assigned to their respective specimen and block, clarifying any
discrepancies in the "reasoning_summary"
```

**SDoc8: Standardize IHC/FISH Template**

```
7. Is there any text segmented under a specimen and block that appears to contain test results for a
different specimen and block?
 - Ensure that the test results are correctly assigned to the correct specimen and block. If there is
any doubt, we should return the reasoning in the reasoning_summary field of the JSON

## Unique Instructions

Apply specific instructions for {{panel}}, if any unique instructions are applicable comment on them
in your reasoning summary
{{standardize_panel_instructions}}

## Task 2, standardize and organize test names and results

Return a JSON with standardized test names and results for each specimen and block
- Each item corresponds to a single test result for a specimen, block and test name
- Format field names as "specimen_SPECIMENNAME_BLOCKNAME_TESTNAME", field values as standardized
results
- The test names and results should be standardized as described in the task 1 instructions
- Attempt regex matching for names and results; use synonyms appropriately
- If there are truly no matches for a test name or result, then the test name and result should be
returned as is, prefixed by 'Other-'
- When a block number is not provided, use 0 as a placeholder
- Remember to follow the naming conventions for result modifiers and patterns.
 - If there are no modifiers, return only the standardized result from the 'status' section. Every
test should have one standardized result from the status section.
 - If there are modifiers, return the standardized terms from the appropriate sections first with a
comma after the status, then the modifiers separated by a space
 - One or more of the modifiers, intensity, extent, and pattern may be present, return them in the
order of intensity, extent, pattern if they are present. If one or more are not present they should be
omitted from the response
- If there are no tests or test results found in the text, return only the reasoning_summary with your
explanation

## Example 1

Example of properly formatted response for a report with 1 specimen, specimen Z, with one block Z1,
and two tests with names TT1 and TT2, the results of which are "Positive" and "Negative" respectively.
In this example pretend that TT1 is in the list of test names provided but TT2 is not, and there are
no good matches nor non trivial synonyms for TT2:
{
"reasoning_summary": "<summary of reasoning for selecting the standardized labels and test results for
each specimen and block. Also how it was determined that which specimen and block test results
correspond to>",
"specimen_Z_block_Z1_TT1": "Positive",
"specimen_Z_block_Z1_Other-TT2": "Negative"
}

## Example 2

A report contains a specimen F with two results for tests RR7 and TT9 from both blocks F5 and F7 and
the result is positive for all tests, but test RR7 has the result modifiers in the text 'greatly
positive in stripe pattern' and a specimen G with a single negative result for test UUT from an
unspecified block. All test names are in the list of names provided, and the test result modifier
'greatly' is in the list of extent test result modifiers, and the result modifier 'striped' is in the
list of pattern modifiers. The JSON would look like this:
{
"reasoning_summary": "<summary of your reasoning>",
"specimen_F_block_F5_RR7": "Positive, greatly striped",
"specimen_F_block_F5_TT9": "Positive",
"specimen_F_block_F7_RR7": "Positive, greatly striped",
"specimen_F_block_F7_TT9": "Positive",
"specimen_G_block_G0_UUT": "Negative"
```

**SDoc8: Standardize IHC/FISH Template**

```
}

## Example 3

Here is an example that incorporates regex matching and synonyms. For the sake of this example, we
will assume that the provided list of test names includes the test "RR7" and "TT9", with "TT9" having
a non trivial synonym "tumor test 9".
The report text contains a single specimen Q and block Q2. In the provided text one of the test names
is spelled "RR-7", and the result is "positive". The second test reported is "tumor test 9" The result
for this test is "neg"
The JSON would look like this, note the use of standardized test names and results:
{
"reasoning_summary": "<summary of your reasoning>",
"specimen_Q_block_Q2_RR7": "Positive",
"specimen_Q_block_Q2_TT9": "Negative"
}

## Example 4

In this example, an outside institution used an unconventional naming convention for a specimen
"specimen 1" and an internal conventional name was NOT provided. The prior LLM has converted the
specimen name to A and the block names to A0 as it is not clear which block was used. The test "p-mas"
was performed with the block not specified and the result is "positive profuse", pretend that the
standardized list contains "P-MAS" and contains under the 'extant' section the extent test result
modifier 'profusely' Then the test "YY9" was performed in house with the results "positive
(reorganized)" Pretend that the standardized list contains "YY9" both the results "Positive" and
"Reorganized" but the unique instructions indicate that the term 'Reorganized' is preferred when BOTH
positive and reorganized are present.
The JSON would look like this, note the use of the internal naming convention, standardized test names
and results, and use of the unique instructions:
{
"reasoning_summary": "<summary of your reasoning>",
"specimen_A_block_A0_P-MAS": "Positive, profusely",
"specimen_A_block_A0_YY9": "Reorganized"
}

## Example 5

In this example, there are TWO specimens from an outside report, and they have non standard names. It
is not clear which specimen was used for the tests and only one set of test results for tests, as
written in the report, "Y-9", "Q5", and "MOP1" with results "intermittently positive in a polka
pattern", "<5% of cells staining" and "Positive, (plastified)" respectively. For this example, let's
pretend "Y-9" is not in the list of test names and does not have any synonyms. The tests "MOP-1" and
"Q5" are in the list. For the "Y-9" result pretend that the results contains "intermittent" under
extent and "polka" under pattern. The "Q5" test is specified in the instructions as having a
percentage result, and there are specific instructions to favor the status "plastified" over
"Positive". The JSON would look like this, note the use of the internal naming convention,
standardized test names and results, and use of the unique instructions, as well as the multiple
specimens with the specimen used not known:
{
"reasoning_summary": "<summary of your reasoning>",
"specimen_X_block_X0_Other-Y-9": "Positive, intermittent polka",
"specimen_X_block_X0_Q5": "<5%",
"specimen_X_block_X0_MOP-1": "Plastified"
}

## Reminders:

It is vital that the entirety of the returned text is a valid JSON

Remember to standardize names and results based on the following lists:
```

##### SDoc8: Standardize IHC/FISH Template

```
When reporting a test name or result, report it standardized to how it appears in the below lists,
even if the capitalization, punctuation, or grammar is different in the text.
{{panel_test_names}}
{{panel_test_results}}

Here is a reminder of the tests that have non trivial synonyms, ensure that you use the standardized
name for these tests:
{{panel_test_synonyms}}
Remember to only use the 'Other-' prefix if there is no fuzzy match for the test name in the list
provided and the test name is not in the list of non trivial synonyms.

# Segmented Text:

Here is the segmented text that you will be working with
{{segment_2_panel_specimen_output}}
```
