## Supplementary figures and images for "Prompts to Table: Specification and Iterative Refinement for Clinical Information Extraction with Large Language Models"

### SFigure 1

#### sFigure 1: Internal Cohort Selection Details

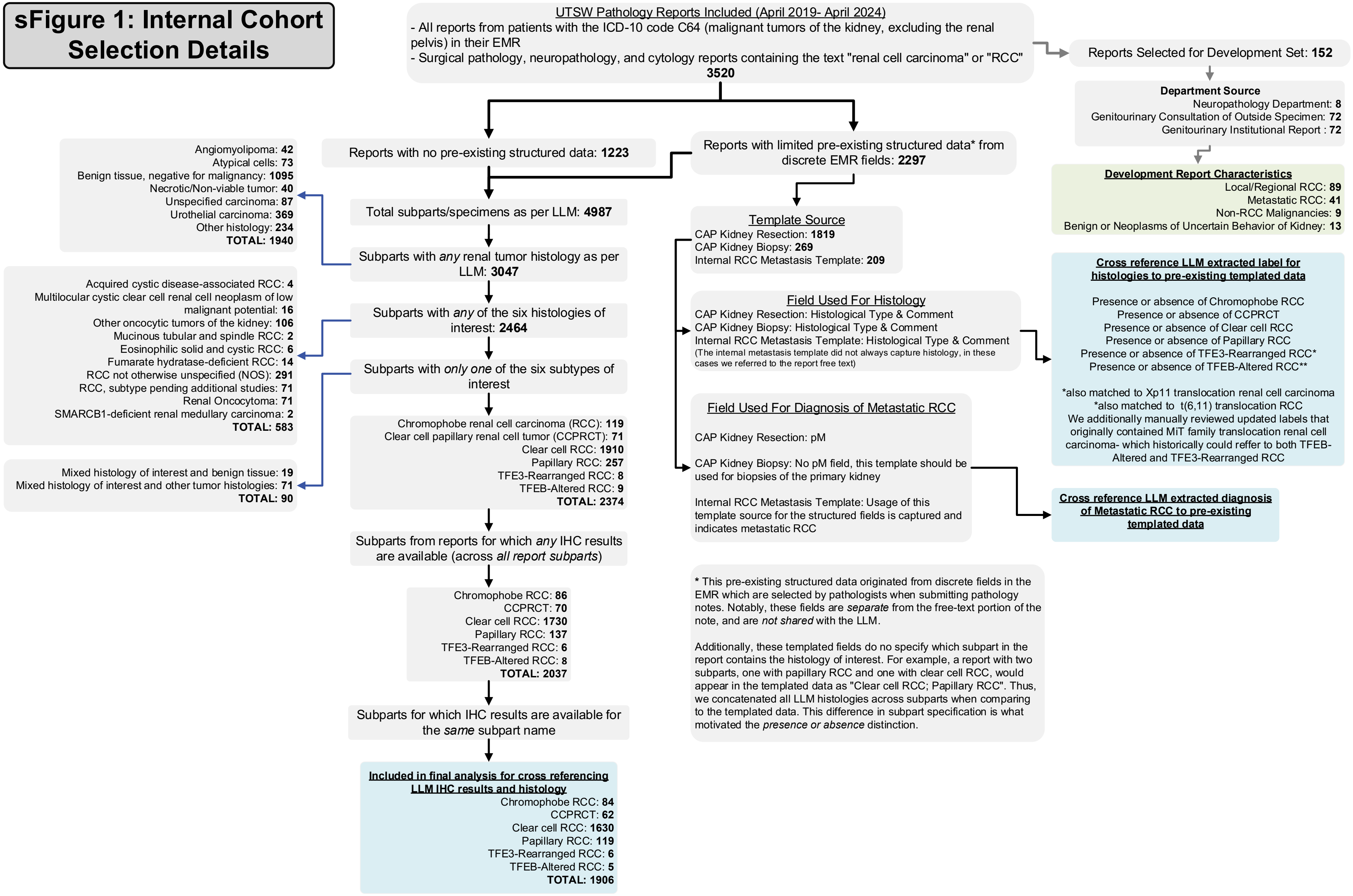
